# Bidirectional Mendelian randomization supports bidirectional causality between telomere length and clonal hematopoiesis of intermediate potential

**DOI:** 10.1101/2021.02.26.21252199

**Authors:** Tetsushi Nakao, Alexander G. Bick, Margaret A. Taub, Seyedeh M. Zekavat, Md M. Uddin, Abhishek Niroula, Cara L. Carty, John Lane, Michael C. Honigberg, Joshua S. Weinstock, Akhil Pampana, Christopher J. Gibson, Gabriel K. Griffin, Shoa L. Clarke, Romit Bhattacharya, Themistocles L. Assimes, Leslie S. Emery, Adrienne M. Stilp, Quenna Wong, Jai Broome, Cecelia A. Laurie, Alyna T. Khan, Albert V. Smith, Thomas W. Blackwell, Zachary T. Yoneda, Juan M. Peralta, Donald W. Bowden, Marguerite R. Irvin, Meher Boorgula, Wei Zhao, Lisa R. Yanek, Kerri L. Wiggins, James E. Hixson, C. Charles Gu, Gina M. Peloso, Dan M. Roden, Muagututi’a S. Reupena, Chii-Min Hwu, Dawn L. DeMeo, Kari E. North, Shannon Kelly, Solomon K. Musani, Joshua C. Bis, Donald M. Lloyd-Jones, Jill M. Johnsen, Michael Preuss, Russell P. Tracy, Patricia A. Peyser, Dandi Qiao, Pinkal Desai, Joanne E. Curran, Barry I. Freedman, Hemant K. Tiwari, Sameer Chavan, Jennifer A. Smith, Nicholas L. Smith, Tanika N. Kelly, Bertha Hildalgo, L. Adrienne Cupples, Daniel E. Weeks, Nicola L. Hawley, Ryan L. Minster, The Samoan Obesity, Lifestyle and Genetic Adaptations Study (OLaGA) Group, Ranjan Deka, Take T. Naseri, Lisa de las Fuentes, Laura M. Raffield, Alanna C. Morrison, Paul S. Vries, Christie M. Ballantyne, Eimear E. Kenny, Stephen S. Rich, Eric A. Whitsel, Michael H. Cho, M. Benjamin Shoemaker, Betty S. Pace, John Blangero, Nicholette D. Palmer, Braxton D. Mitchell, Alan R. Shuldiner, Kathleen C. Barnes, Susan Redline, Sharon L.R. Kardia, Gonçalo R. Abecasis, Lewis C. Becker, Susan R. Heckbert, Jiang He, Wendy Post, Donna K. Arnett, Ramachandran S. Vasan, Dawood Darbar, Scott T. Weiss, Stephen T. McGarvey, Mariza de Andrade, Yii-Der Ida Chen, Robert C. Kaplan, Deborah A. Meyers, Brian S. Custer, Adolfo Correa, Bruce M. Psaty, Myriam Fornage, JoAnn E. Manson, Eric Boerwinkle, Barbara A. Konkle, Ruth J.F. Loos, Jerome I. Rotter, Edwin K. Silverman, Charles Kooperberg, Siddhartha Jaiswal, Peter Libby, Patrick T. Ellinor, Nathan Pankratz, Benjamin L. Ebert, Alexander P. Reiner, Rasika A. Mathias, Ron Do, NHLBI Trans-Omics for Precision Medicine (TOPMed) Consortium, Pradeep Natarajan

## Abstract

Human genetic studies support an inverse causal relationship between leukocyte telomere length (LTL) and coronary artery disease (CAD), but directionally mixed effects for LTL and diverse malignancies. Clonal hematopoiesis of indeterminate potential (CHIP), characterized by expansion of hematopoietic cells bearing leukemogenic mutations, predisposes both hematologic malignancy and CAD. *TERT* (which encodes telomerase reverse transcriptase) is the most significantly associated germline locus for CHIP in genome-wide association studies. Here, we investigated the relationship between CHIP, LTL, and CAD in Trans-Omics for Precision Medicine (TOPMed) program (N=63,302) and UK Biobank (N=48,658). Bidirectional Mendelian randomization studies were consistent with LTL lengthening increasing propensity to develop CHIP, but CHIP then in turn hastening LTL shortening. We also demonstrated evidence of modest mediation between CHIP and CAD by LTL. Our data promote an understanding of potential causal relationships across CHIP and LTL toward prevention of CAD.

## Introduction

Telomeres consist of repetitive DNA sequences with associated protective proteins,^1^ that stabilize chromosomes by several mechanisms.^2^ Shortening of telomeres during successive mitoses aims to protect the remaining chromosomal DNA. Reverse transcription by the telomerase complex mitigates telomere attrition in cells requiring frequent division such as hematopoietic stem cells.^3-5^ However, with aging, telomeres continue to shorten and protective mechanisms are less efficient leading to accumulating senescent cells with shortened telomeres providing a fertile substrate for genomic instability.^6^ Senescent cells also acquire a proinflammatory senescence-associated secretory phenotype (SASP), which promotes aging-related cardiovascular disease.^7^

While Mendelian randomization (MR) studies consistently support an inverse causal relationship between leukocyte telomere length (LTL) and coronary artery disease (CAD), the relationship between LTL and cancer is more complex.^8-10^ *In vitro* studies indicate that short telomeres promote genomic instability, thereby leading to malignancies,^2,11^ and most tumor cells have shortened telomeres.^12-15^ Mendelian disorders characterized by severe telomere shortenings, such as aplastic anemia and dyskeratosis congenita, are characterized by premature aging, organ damage, and high rates of malignant blood disorders.^16,17^ However, MR studies indicate that longer LTL may be causally associated with an increased incidence of various malignancies, such as lung adenocarcinoma, glioma, melanoma, or leukemia.^8-10,18-21^

Age-related clonal hematopoiesis of indeterminate potential (CHIP), characterized by clonally expanded hematopoietic cells bearing leukemogenic mutations (most commonly in *DNMT3A, TET2, ASXL1*, and *JAK2*) without clinical hematologic disorders, represents a pre-malignant condition. CHIP strongly predicts future risk for myeloid malignancy and human and murine data indicate that CHIP is a causal risk factor for CAD as well.^22-27^ In cross-sectional analyses, the presence of CHIP correlates with shorter LTL adjusting for age.^28^ However similar to aforementioned cancer studies, in genome-wide association analyses of CHIP, the most significant risk allele resides in the *TERT* locus^29^ (encoding telomerase reverse transcriptase) and is associated with lengthened LTL.^30^ Whether and how CHIP and LTL are causally related is unknown, and whether this relationship influences CHIP-associated risk for CAD is unknown.

Here, we investigated the relationships between LTL, CHIP, and CAD to address these questions using LTL estimated from whole exome sequencing (WES) data in the UK Biobank (N = 48,658) and from whole genome sequencing (WGS) data in the NHLBI TOPMed program (N = 63,302).^30,31^

After estimating the associations across LTL, CHIP, and CAD, we assessed these associations for evidence supporting bidirectional causality using MR. Finally, we estimated the mediation effect of LTL for the CHIP-associated CAD risk.

## Results

### Baseline characteristics

CHIP was detected, and LTL was estimated with blood DNA-derived WGS from the U.S. National Heart, Lung, and Blood Institute (NHLBI) Trans-Omics for Precision Medicine (TOPMed) program^32^ and WES from the UK Biobank.^33^ In TOPMed, CHIP calls and LTL estimates were obtained from whole genome sequence analyses previously published.^29,30^ In TOPMed, after excluding kinship through second-degree relatives, 63,302 individuals had both indices measured of whom 36,507 (57.7 %) were female. The mean age was 54.3 years old (standard deviation (SD) 18.0) at the time of blood draw, and 29,171 (64.6%) were of European ancestry (Fig. 1, Supplementary Fig. 1, Supplementary Table 1). CHIP calls from the first WES samples released from UK Biobank were obtained^27,33^ with some modifications (see Methods), yielding 48,658 individuals after excluding related individuals (second-degree) and discordance between genetic sex and self-reported sex. Among the UK Biobank participants included, 26,515 (54.5 %) were female. The mean age was 56.5 years old (SD 8.0), and 43,227 (93.9%) were of European ancestry. In total, 3,284 (TOPMed: 5.1 %) and 2,273 (UK Biobank: 4.6 %) individuals had evidence of at least one CHIP-related mutation. Of these, 2,862 (TOPMed: 4.5 %) and 1,044 (UK Biobank: 2.1 %) individuals had variant allele frequency greater than 0.10, a threshold previously associated with increased CAD incidence^24,25,27^ (Supplementary Table 1).

**Fig. 1:**
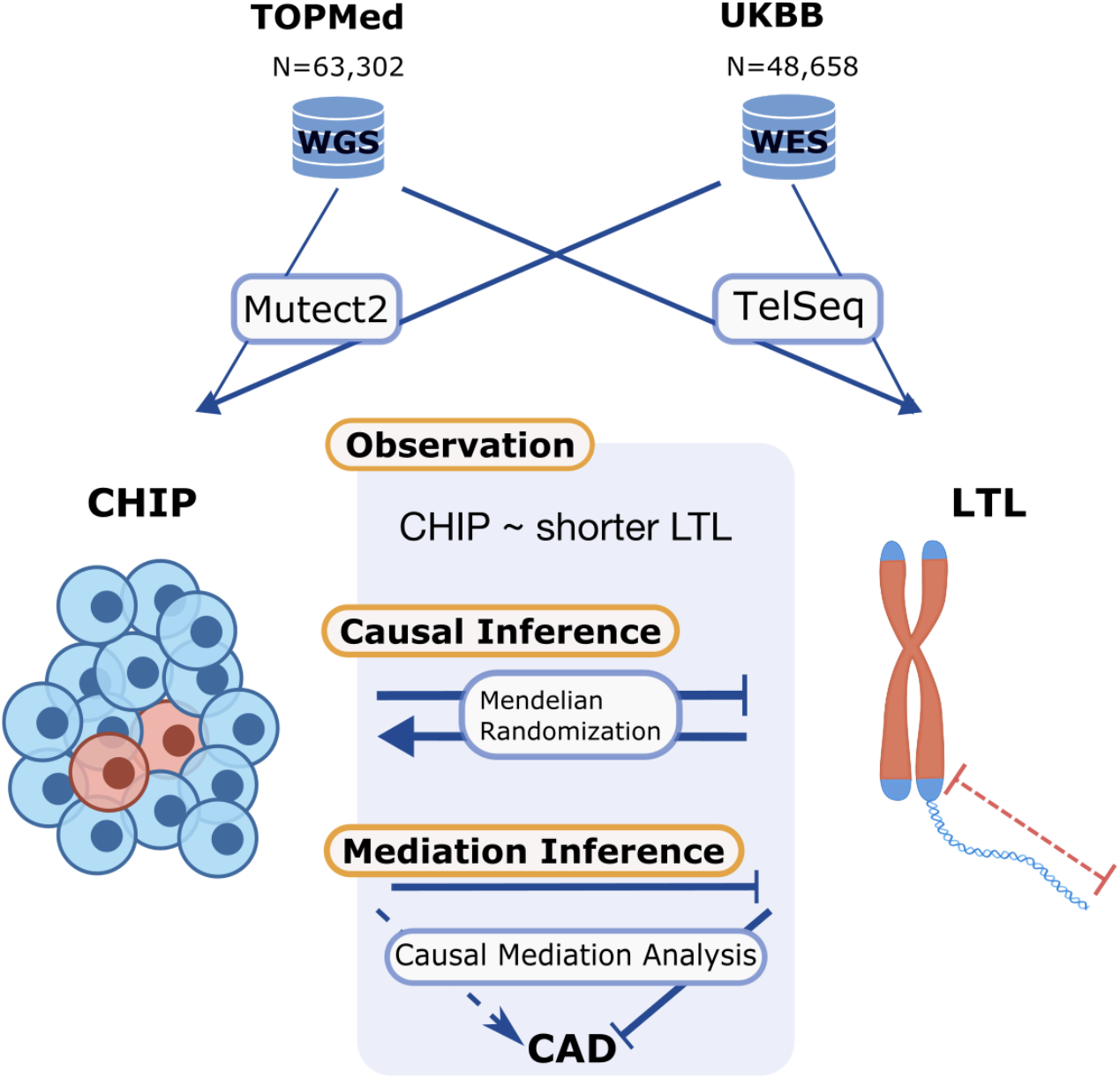
Analytical procedure in this study. TOPMed (N=63,302) and UK Biobank (N=48,658) are used as the study cohorts. CHIP associated mutations are detected by Mutect2. Telomere length was estimated by TelSeq. Observational study and causal inference by bidirectional Mendelian randomization were performed between LTL and CHIP. CHIP was associated with shorter LTL. Germline genetic factors that increase CHIP development were associated with shorter LTL, whereas germline genetic factors that increase LTL were associated with developing CHIP. Mediation effect of LTL on CHIP related CAD risk increment was detected by causal mediation analysis. CHIP: clonal hematopoiesis of indeterminate potential, LTL: leukocyte telomere length, TOPMed: Trans-Omics for Precision Medicine, UKBB: UK Biobank.

### Telomere length estimation using WES in UK biobank

The TelSeq algorithm was applied to the 49,739 CRAM files of WES in the UK Biobank to estimate LTL. The same algorithm was used to estimate LTL from WGS data in TOPMed.^30^ Consistent with restricted sequencing inherent to WES, the estimated absolute LTL (mean ± SD (kb): 0.07 ± 0.08) from WES in UK Biobank was much shorter than that estimated from WGS in TOPMed (mean ± SD (kb): 3.27 ± 1.01) or from conventional LTL measured by Southern blot (mean ± SD (kb): 6.87 ± 0.62) measured in a subset of the Women’s Health Initiative (WHI) (N = 686) (Supplementary Fig. 2a,b). LTL estimation was batch-corrected with the first nine principal components (PCs) generated by NGS-PCA (https://github.com/PankratzLab/NGS-PCA) which uses calculated from read coverage information that is standardized to assess the relative value (Supplementary Fig. 2c). Despite much shorter absolute estimates, the batch-corrected LTL normalized distribution much more closely approximated the expectation and so we focus on LTL SD unit hereafter. Variants previously associated with LTL were associated with LTL in UK Biobank similarly to prior qPCR-based telomere estimation (T/S ratio),^10^ but with relative effect deflation (R^2^ = 0.69, *P* = 3.02×10^−6^) (Supplementary Fig. 3). Age was correlated with LTL in both TOPMed WGS (β = -0.023, *P* < 2×10^−16^) and UK Biobank WES (β = -0.0098, *P* < 2×10^−16^) after adjustment with sex, ever smoking, first 11 genetic PCs (Supplementary Fig. 2d).

### Shorter LTL is associated with increased CHIP prevalence and increased CAD incidence

We performed association studies between LTL, CHIP, and CAD separately in TOPMed and UK Biobank, followed by meta-analyses. Consistent with prior reports,^4^ CHIP with variant allele frequency (VAF) > 0.10 was associated with shorter LTL in meta-analysis results after adjustment for age, sex, ever smoking, body mass index (BMI), study, sequencing center, and first 11 genetic PCs (β = -0.13; 95% CI -0.16:-0.096; P(heterogeneity)=0.02) (Fig. 2a, Supplementary Fig. 4a). Here, VAF was defined as the largest clone if multiple CHIP clones were detected in the same individual. CHIP with VAF < 0.10 was associated with shorter LTL in TOPMed (WGS) but not in UK Biobank (WES), likely due to reduced sensitivity of smaller CHIP clones and improved LTL estimation with WGS relative to WES (Supplementary Fig. 4b).

**Fig. 2:**
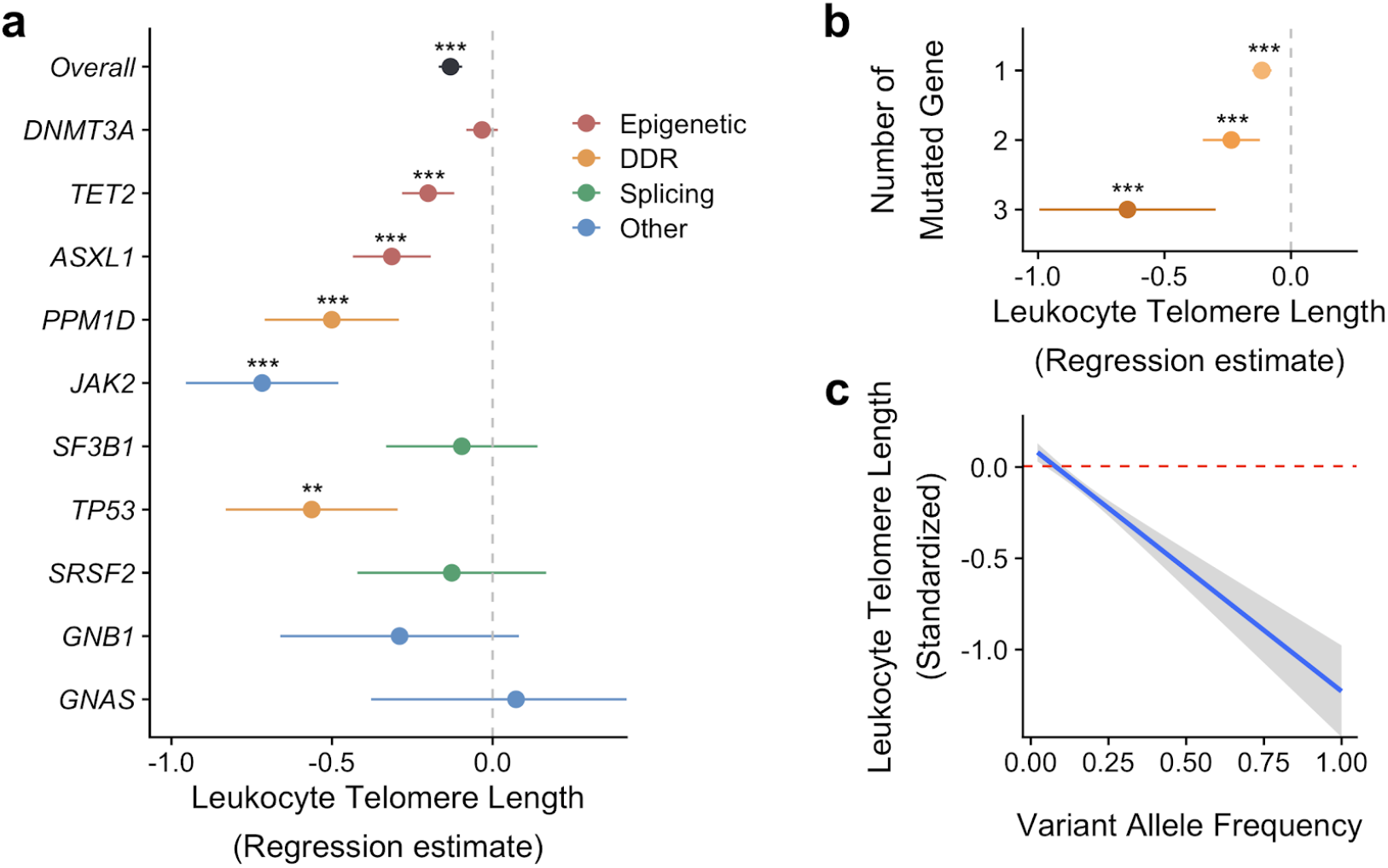
CHIP prevalence and VAF are associated with shorter LTL. The associations of CHIP with LTL were assessed by linear regression model both in TOPMed and UK Biobank, then meta-analyzed by fixed effect model. Both models were adjusted with age, sex, ever smoking, body mass index, first 11 genetic principal components, study within TOPMed and sequencing center (study and sequencing center are only applicable to TOPMed). The prevalence of CHIP with greater than 10% VAF associations were evaluated for overall and each mutated gene (**a**) and for each number of mutated genes in the same individuals (**b**). **c** The correlation between LTL and VAF among the population with CHIP from both TOPMed and UK Biobank pooled analysis is displayed. A subset in TOPMed with age 40 to 70 was included in the analysis to align with the age distribution in UK Biobank. Red dotted line represents the average LTL in the population without CHIP. ** *P* < 0.01, *** *P* < 0.001, after Bonferroni’s correction if applicable. CHIP: clonal hematopoiesis of indeterminate potential, DDR: DNA damage repair, LTL: leukocyte telomere length, TOPMed: Trans-Omics for Precision Medicine, VAF: variant allele frequency.

Prior cell-based studies have shown that *DNMT3A* loss of function increases telomere length,^34^ *TET2* loss of function decreases telomere length;^35,36^ and *p53* (TP53) protects telomeres from DNA damage.^37^ Thus, we estimated the effect size of each mutated gene on LTL (Fig. 2a, Supplementary Fig. 5, Supplementary Table 2). *DNMT3A* did not show a significant association with LTL, whereas *TET2, ASXL1, PPM1D, JAK2*, and *TP53* were significantly associated with shorter LTL. Multiple CHIP mutations in the same individuals had an additive effect on shorter LTL (Fig. 2b, Supplementary Fig. 6a). Each additional CHIP-related mutation yielded an effect size of -0.11 when meta-analyzed across both cohorts (95% CI -0.14:-0.086; *P* = 6. 83×10^−14^) (Supplementary Fig. 6b).

Among those with CHIP, increasing VAF correlated strongly with shorter LTL after adjustment in both cohorts (β = -1.12 / 1% of VAF; 95% CI -1.40:-0.84; P(heterogeneity)=0.14) (Fig. 2c, Supplementary Fig. 7).

We next assessed the association of LTL with CAD using subsets of the cohorts with information on incident CAD in TOPMed (N = 27,937) and in UK Biobank (N = 48,658). Individuals who experienced CAD prior to the blood draw used to determine CHIP status were excluded from TOPMed (N = 416) and UK Biobank (N = 1,357) analyses and follow up duration was calculated starting the time at the blood draw. Incident CAD was defined both in TOPMed and UK Biobank by ischemic heart disease, including myocardial infarction, and coronary revascularization (Supplementary Table 3).

We used Cox proportional hazard models to evaluate the association between LTL and CAD including multivariable adjustment with covariates age, sex, ever smoking, BMI, hypercholesterolemia, the first 11 genetic PCs, study within TOPMed, and sequencing center (study and sequencing center were only applicable to TOPMed). Missing covariates excluded 8,324 TOPMed (8,200 COPDGene participants without blood lipids, and 124 other individuals across TOPMed missing covariates) and 1,224 UK Biobank individuals from the analysis (Supplementary Fig. 1). Of the remaining 19,176 TOPMed and 46,077 UK Biobank individuals, 3,283 TOPMed (17.1%) and 1,379 UK Biobank (3.0%) participants developed CAD during the follow-up duration (mean (SD) duration 12.0 (5.8) years in TOPMed and 10.0 (1.5) years in UK Biobank) (The number of individuals included in the final analysis in each TOPMed cohort was shown in Supplementary Table 4). Shorter LTL was associated with increased CAD risk (Hazard Ratio = 1.07; 95% CI 1.04:1.10; *P* (heterogeneity) = 0.71) (Supplementary Fig. 8) as previously reported.^38-40^

### Mendelian randomization studies indicate that CHIP causes shortened telomeres

We performed one-sample MR for CHIP on LTL with the TOPMed cohort. Instrumental variables (IVs) were utilized from a previously reported GWAS of CHIP.^29^ To avoid the possible bias from reverse causality, we filtered discovered IVs using the Steiger test that identified the *TERT* locus as having a significantly higher correlation with LTL than CHIP (*P* = 0.0105); hence, SNPs at *KPNA4/TRIM59* and *TET2* loci were valid IVs (Supplementary Table 5). The significance of each variant supported the robust association with exposure, the first assumption of Mendelian randomization (Supplementary Table 5). The MR analysis with 2 IVs was consistent with an inverse causal effect of CHIP on LTL (Estimate = -0.81; 95% CI -1.40:-0.23; *P* = 0.0063) (Fig. 3). No statistical evidence of endogeneity for IVs used was shown by the Sargan test (*P* = 0.306). Single IV analysis demonstrated consistent effect sizes across 2 IVs (Supplementary Fig. 9).

**Fig. 3:**
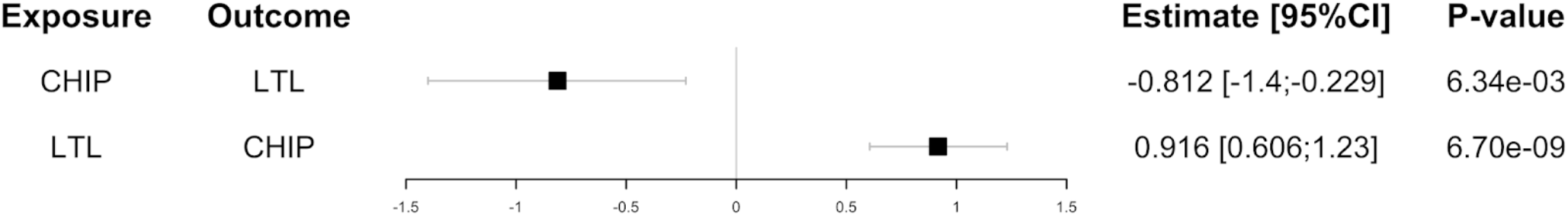
Bidirectional Mendelian randomization studies showed the negative causal effect of CHIP on LTL and the positive causal effect of LTL on CHIP. Bidirectional one-sample MR was performed to assess the causal effect of both CHIP on LTL and LTL on CHIP. TOPMed was used for IV discovery for CHIP on LTL, Li et al *AJHG* 2020 for LTL on CHIP. IVs were clumped if < 10 Mb apart and in linkage disequilibrium (R^2^ > 0.001 calculated in European ancestry from 1000 Genome project). IVs were further assessed by Steiger test to mitigate the effect of reverse causation resulting in 2 and 16 IVs, respectively. TOPMed was used as the test cohort for both CHIP on LTL and LTL on CHIP. Used IVs and cohorts for each analysis are summarized in Supplementary Tables 5 and 7. CHIP: Clonal hematopoiesis of intermediate potential, CI: confidence interval, IV: Instrumental variable, LTL: Leukocyte telomere length, MR: Mendelian randomization, TOPMed: Trans-Omics for Precision Medicine, UKBB: UK Biobank.

We next performed a two-sample MR study. The subset of European ancestry participants from TOPMed was used as the CHIP exposure cohort and publicly available summary statistics for LTL from the ENGAGE (European Network for Genetic and Genomic Epidemiology, N = 37,684)^41^ consortium were used for the outcome. The low heritability for CHIP, limitation to European ancestry samples, and removal of the one significant (*P* < 5×10^−8^) locus (*TERT* locus) in a European-only CHIP GWAS^29^ due to Steiger filtering (*P* = 0.011), to minimize bias from potential reverse causality, limited the power for conventional two-sample MR analyses. Therefore, we used MR-RAPS^42^ to accommodate many weak IVs with a higher *P*-value threshold. Sixty IVs were used with *P* < 10^−4^ significance threshold and SNPs were clumped if R^2^ > 0.001 or located within 10 Mb from each other. The *TERT* locus was excluded from both Steiger filtering and Cochran’s Q test, and one non-inferable palindromic SNP was excluded (Supplementary Table 6). The remaining fifty-eight SNPs were used as IVs and supported an inverse causal association of CHIP on LTL (β = -0.017, *P* = 0.04). Leave-one-out analysis showed the overlapping locus with one-sample MR analysis (*KPNA4/TRIM59*, rs4679885) is not a significant factor accounting for the causal inference (Supplementary Fig. 10).

### Mendelian randomization studies indicate that longer LTL cause CHIP acquisition

We performed one-sample MR using TOPMed individual-level data from LTL toward CHIP. IVs were discovered from GWAS of LTL conducted in an independent cohort^10^ and pruned as 10 Mb apart and in linkage disequilibrium. Two-stage least square regression showed the causal association of LTL on all CHIP (Estimate = 0.92, 95% CI 0.61:1.23; *P* = 6.7×10^−9^) (Fig. 3). Though Sargan test indicated endogeneity of used IVs (*P* < 2.2×10^−16^), we do not have an appropriate strategy to find pleiotropy in one-sample MR. Hence, the analysis was re-examined after exclusion of outliers detected by MR-PRESSO in the following replication analysis. Outlier exclusion still demonstrated significant causal association (Estimate = 0.48; 95% CI 0.13:0.83; *P* = 0.0062) without statistically significant evidence of pleiotropy (Sargan test; *P* = 0.83).

A replication analysis was performed by two-sample MR studies. Previous LTL summary statistics^10^ were used as the exposure and the white British subset of the UK Biobank was used as the CHIP outcome cohort (N = 42,201), which are both independent from TOPMed used in one-sample MR. The positive potential causal effect of LTL on CHIP was shown in the conventional two-sample MR approach (Inverse Variance Weighted (IVW) method; Estimate = 1.06; 95% CI 0.39:1.74; *P* = 1.89×10^−3^) (Supplementary Fig. 11). The global test by MR-PRESSO^43^ suggested significant horizontal pleiotropy before outlier exclusion (*P* < 1.0×10^−4^) and detected the *TERT* and *ATM* loci as outliers (Supplementary Table 7). While the leave-one-out analysis showed the *TERT* locus variant had the most significant effect among IVs, the analysis remained robust (Supplementary Fig. 12). Outlier exclusion supported a significant causal association of LTL on CHIP by MR-PRESSO in two-sample MR (Estimate = 0.79; 95% CI 0.24:1.34; *P* = 0.014) (Supplementary Fig. 11) without significant statistical evidence of horizontal pleiotropy (Global Test: *P* = 0.15). Re-evaluation of all the models after outlier exclusion showed stable estimates across methods indicating robust causal inference (Supplementary Fig. 13).

We next evaluated the relationship of LTLs with the occurrence of acquired genome-wide singleton single nucleotide substitution. Using WGS from a subset of the TOPMed study population (N = 28,392), we tabulated per-individual genome-wide somatic mutations. Outlier-excluded 14 IVs discovered in the previous section were used for MR study. MR analyses supported a causal relationship between longer LTL and increased somatic mutations in one-sample MR study (Supplementary Fig. 14). Next, we assessed the effect of LTL for COSMIC signature version 2 (https://cancer.sanger. ac.uk/cosmic/signatures_v2). Failure of DNA double-strand break repair by homologous recombination (Signature 3) and other signatures with unknown etiologies (Signatures 17, 28, and 30) associate with longer LTL in MR study (Supplementary Fig. 15). These observations suggested that longer LTL promotes CHIP acquisition by accelerating mutagenesis. The *TERT* locus variant associated prominently with mutational occurrence in line with the pleiotropic effect detected in MR studies (Supplementary Fig. 16).

### Causal Mediation analysis of LTL for CHIP associated CAD risks

We assessed the mediation effect of LTL on CHIP-associated CAD risk in UK Biobank. The proportion of mediation effect of LTL in the total effect of CHIP to CAD was estimated as 1.47 % (95% CI 0.18:5.3 %; *P* = 0.02) using “*mediation*” package^44^ in R (R Foundation for Statistical Computing, Vienna, Austria) (Table 1). Both mediator and outcome models were adjusted for age, sex, ever smoking, previous type 2 diabetes mellitus, previous hypercholesterolemia, previous hypertension, and the first 11 genetic PCs. We performed a replication analysis using the Women’s Health Initiative (WHI) cohort subset of the TOPMed cohort (N = 3,734), since sufficient covariates information to adjust models precisely were available. CAD was defined as the composite of myocardial infarction and coronary revascularization. The proportion of causal mediation effect of LTL in the total effect of CHIP to CAD was estimated as 5.2 % (95% CI 0. 10:33 %; *P* = 0.02) in WHI.

**Table 1:** Causal mediation analysis showed mediation effect of LTL for CHIP associated CAD risk. The mediation effect of LTL for CHIP associated CAD risk were estimated by “mediation” package in R. Mediation effect of 0 indicates that LTL does not mediate the CHIP associated CAD risks, and mediation effect of 1 indicate that LTL mediates all of the CHIP related CAD risks. The *P*-value reflects whether the proportion of the mediation effect on the CHIP related CAD risks is 0 vs. not 0. Both mediator and outcome models are adjusted by age, sex, ever smoking, prevalent type 2 diabetes, prevalent hypercholesterolemia, prevalent hypertension, and the first 11 genetic principal components in UK Biobank, and age at blood draw, ever smoking, race, dyslipidemia, hypertension, body mass index, WHI inverse probability weight (to account for the non-random selection of women for whole genome sequencing in WHI), history of hormone therapy, history of hysterectomy, and first 11 genetic principal components in WHI. CAD: coronary artery disease, CHIP: clonal hematopoiesis of intermediate potential, LTL: leukocyte telomere length, WHI: Women’s Health Initiative.

|  | **Proportion of Mediation Effect of LTL**<br>**For CHIP Associated CAD Risk (95% CI)** | ***P*-value** |
| --- | --- | --- |
| **UK Biobank**<br>**(N = 44,921)** | 0.014<br>(0.0018-0.053) | 0.02 |
| **WHI**<br>**(N = 3,734)** | 0.052<br>(0.010-0.33) | 0.02 |

## Discussion

We used observational and Mendelian randomization studies to examine how processes regulating LTL and CHIP acquisition interrelate, and how they influence CAD risk. Consistent with prior observational epidemiologic analyses, CHIP and LTL were inversely correlated. Bidirectional Mendelian randomization supported the hypotheses that longer LTL promotes CHIP acquisition whereas CHIP in turn shortens LTL potentially in affected cells. While both CHIP and shorter LTL have been independently associated with CAD, causal mediation analysis indicated that a modest fraction of CHIP-associated CAD risk may be mediated by resultant LTL shortening.

Our findings have several implications for the understanding of CHIP, LTL, and CAD. First, we observe a bidirectional causal relationship between LTL and CHIP, advancing our understanding of the malignancy-telomere length association. As described earlier, prior studies have shown complex relationships between LTL and cancer risk.^21^ Several models were proposed, including a heterogeneous multi-hit theory^45^ and the biphasic effect of *TERT* promoter mutation throughout tumor development.^14^ CHIP provides an opportunity to focus on an incipient step of malignant cell development. Our results suggest that longer LTL may promote CHIP acquisition through increasing mutagenesis. One potential model could be that longer telomeres support the longevity of the cells, thus augmenting opportunities to acquire somatic mutations over time, while telomeres begin accelerated shortening once the cell cycle accelerates due to driver mutation acquisition (Fig. 4). Consistent with this model, we observed that increased clone size, a readout of increased cellular cycles, is correlated with shorter LTL.^46^ In the setting of shortened telomere Mendelian syndromes, shortened telomeres promote genomic instability and subsequent acquisition and retention of neoplastic driver mutations.^47^ This may be consistent with the observation that CHIP-associated LTL shortening may hasten subsequent malignancy (Supplementary Fig. 17). Further assessment of longitudinal LTL followup among CHIP positive population would be desired.

**Fig. 4:**
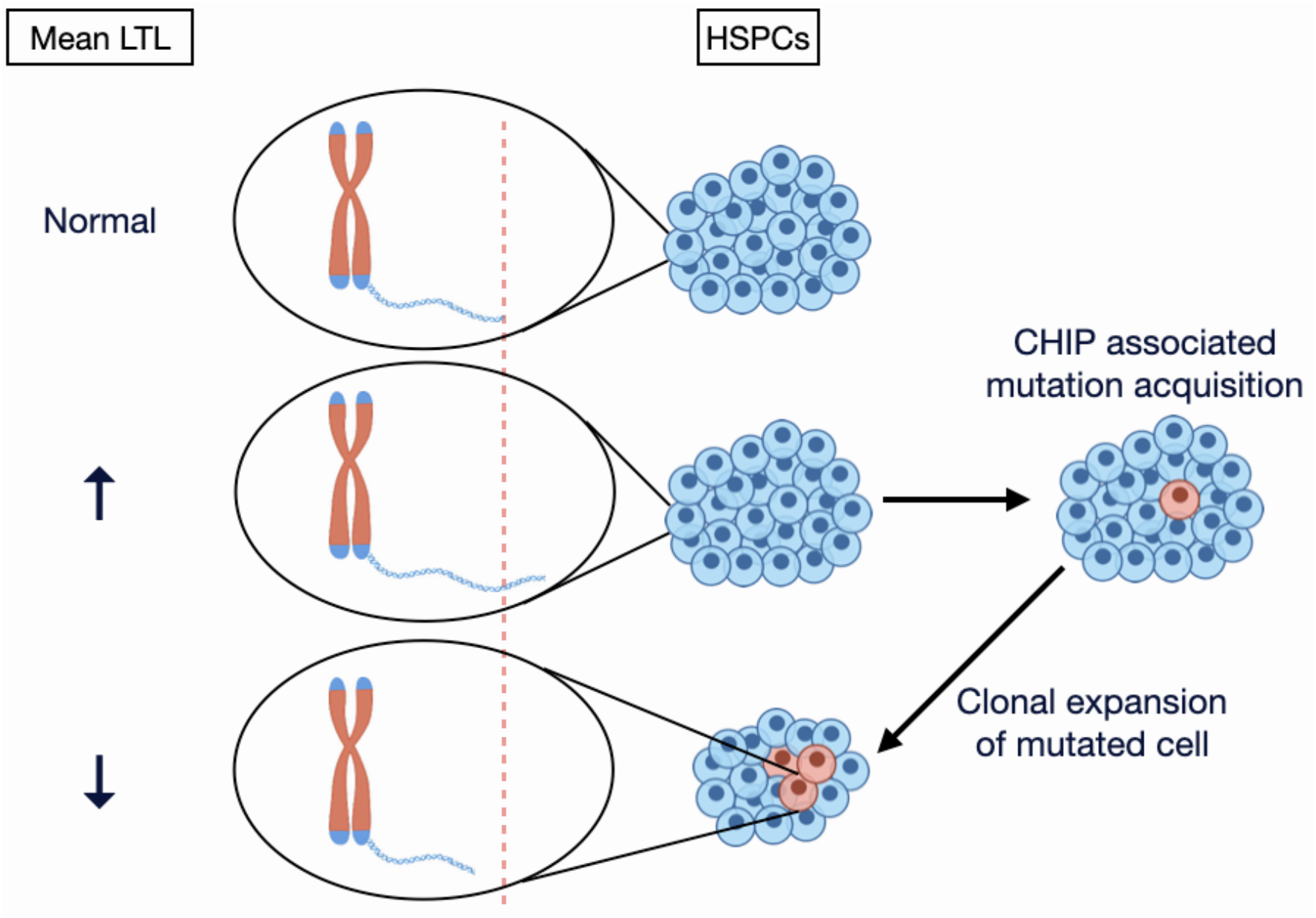
Proposed model of “Telomere Paradox” in CHIP. People with longer mean leukocyte telomere length (LTL) have higher incidence of mutagenesis so that have higher chance to acquire CHIP associated mutations (middle). The cells that acquired CHIP have paradoxically shorter telomeres so that mean LTL decreases as the clone expands (bottom). CHIP: clonal hematopoiesis of intermediate potential, LTL: leukocyte telomere length, HSPC: hematopoietic stem cell.

Second, CHIP-associated CAD risk may be partly attributed to subsequent LTL shortening. Prior cell-based, murine, and human genetic analyses have prioritized the NLRP3 inflammasome pathway in CHIP-associated CAD risk.^24-27,29,48,49^ In the present work, we orthogonally implicate LTL in both the genesis of CHIP, a new CAD risk factor, and its clinical consequences. Our study is underpowered for gene-specific analyses but notably did not observe an association between *DNMT3A* CHIP and LTL alteration. Consistent with this observation, prior work suggests that hematopoietic stem cell loss of *Tet2* leads to shortened telomeres, whereas loss of *Dnmt3a* leads to telomere preservation.^35,50^ Such differences may also partly explain gene-specific differences in CAD risk.^25-27^ While interrupting CHIP-mediated LTL shortening may be a viable strategy to mitigate CHIP-associated CAD risk, this general strategy may be limited to the overall modest estimated mediating effect. However, given heterogeneity observed, this strategy may be more efficiently applied to non-*DNMT3A* CHIP.

Key limitations must be considered in the interpretation of our study findings. First, limited CHIP GWAS availability prevented conventional two-sample MR approaches for CHIP on LTL. The ongoing effort of accumulating CHIP cases would address this issue. Second, the cross-sectional observational nature of our analyses limits inference regarding causal temporal relationships. We employ several sensitivity analyses for Mendelian randomization for robustness. Longitudinal analyses of LTL, CHIP, and incident diseases as well as experimental models are needed to confirm our hypotheses. Third, the causal mediation effect estimate of LTL for CHIP-associated CAD may be limited by conflicting bidirectional causal effect.

In conclusion, we showed a bidirectional relationship between LTL and CHIP, shedding light on the mechanisms by which telomere length contributes to age-related disorders. The causal mediation effect of LTL on CHIP-related CAD incidence suggests the plausibility of developing harmonized therapies for both blood cancer and cardiovascular diseases.

## Data Availability

Data from TOPMed are available via dbGaP.
Data from UK Biobank will be made available through an application.

