## Supplementary Materials for "Bidirectional Mendelian randomization supports bidirectional causality between telomere length and clonal hematopoiesis of intermediate potential"

### Methods

*Study populations*

The UK Biobank is a population-based cohort of >500,000 UK adult residents recruited between 2006-2010 and followed prospectively via linkage to national health records.[33] At the baseline study visit, participants underwent phlebotomy and provided detailed information about medical history and medication use. In the present study, the UK Biobank cohort included adults aged 40-70 years at blood draw with available whole-exome sequence (WES). Follow-up in the UK Biobank occurred through March 2020 for inpatient diagnosis. We excluded samples with consent retraction, excess heterogeneity or missingness in genotyping array, discordance between reported and genetically imputed sex, and excess kinship within second-degree determined by *KING* coefficient > 0.0884 (Supplementary Fig. 1).

From NHLBI's Trans Omics for Precision Medicine (TOPMed) program, whole-genome sequence (WGS) of blood DNA performed in participating studies were used for clonal hematopoiesis of intermediate potential (CHIP) detection and leukocyte telomere length (LTL) estimation in recent studies.[29,32] Comprising studies are largely observational cohorts and have previously been described in detail. Cohorts included in this study are reported in Supplementary Table 4. We excluded samples which had conflicting information for sex and excess kinship within second-degree determined by *KING* coefficient > 0.0884 (Supplementary Fig. 1).

Kinship inference and genetic principal components (PCs) were centrally calculated in both cohorts.

For the two-sample Mendelian randomization (MR) studies, the European ancestry (N = 29,373 in TOPMed, N = 42,201 in UK Biobank) subset was used to avoid bias from population structures in both cohorts.

Since the Women's Health Initiative (WHI) represents one of the largest TOPMed cohorts and had exposure, sufficient covariates, and outcome data available, we used WHI for the replication of mediation analysis for LTL on CHIP associated coronary artery disease (CAD) risks. Briefly, the WHI is a prospective study of women recruited at 40 centers throughout the United States between 1993 and 1998.[51] Participants enrolled in the clinical trial(s) (of hormone therapy, calcium/vitamin D supplementation, and/or dietary modification) or the observational study. Unrelated women in WHI who underwent >30X WGS using blood drawn at age 50-70 years as part of the TOPMed program were included in the present analysis. To avoid the effects of the study intervention on outcomes, we excluded women in the WHI hormone therapy trial with blood draw ≥2 years after the screening visit (N = 483) as previously described.[52]

*Sequencing and CHIP calling*

UK Biobank WES of whole blood-derived DNA was performed using Illumina NovaSeq 6000 platform at the Regeneron Sequencing Center (Tarrytown, NY) as described previously.[33] TOPMed WGS to an average depth of 38x was performed using whole blood-derived DNA, PCR-free library construction, and Illumina HiSeq X technology as described elsewhere.[32]

CHIP mutations were called previously in TOPMed[29] and UK Biobank.[27] CHIP mutations were re-evaluated after the error corrected release of WES in UK Biobank at June 2020. Briefly,

CHIP mutations were detected with GATK MuTect2 software[53] with parameters as previously described.[29,54] Samples were annotated as having CHIP if Mutect2 identifies one or more of a pre-specified list of pathogenic somatic variants.[24,25] Common germline variants and sequencing artifacts were excluded as before. Each study includes both the presence of (1) any CHIP and (2) CHIP with variant allele frequency (VAF) > 0.1, as larger CHIP clones above this threshold have previously been more strongly associated with adverse clinical outcomes.[25,27]

*Estimating LTL from Next-Generation Sequencing of Blood-derived DNA*

The estimation of mean LTL was performed using TelSeq[31] previously in TOPMed with WGS.[30]

Given imperfect capture, TelSeq is expected to be able to estimate LTL from WES.[31] We applied an analogous method to WES in UK Biobank with k = 7, while k = 12 was used for TOPMed WGS. Read coverage was calculated by Mosdepth[55] and principal component analysis was conducted for read coverage. Estimated LTL was log-transformed and linear regression was performed using the first nine PCs. Standardized residuals were used as the relative value of estimated LTL within each study for mean = 0 and standard deviation =1 for each cohort.

*Telomere measurement by Southern blotting*

We measured LTL in a subset of WHI by Southern blotting for direct comparison with TelSeq using WGS. DNA was extracted from baseline (or year 1) blood samples by the 5-prime method (5 PRIME, Inc.; Gaithersburg, MD) and sent in batches over a 1-year period to the Center of Human Development and Aging laboratory at Rutgers for LTL measurement. The laboratory conducting the LTL measurements was blinded to all characteristics of participants.

DNA integrity was assessed visually after ethidium bromide-stained 1% agarose gel electrophoresis (200 V for 2 hours). We required DNA to appear as a single compact crown-shaped band that migrated in parallel with the other samples on the gel. Telomere length in kilobases (kb) was measured by the mean length terminal restriction fragments using the Southern blot method as previously described.[56] Each sample was run in duplicate on different gels and mean LTL was used for statistical analyses. The average inter-assay coefficient of variation for blinded pair sets was 2.0%. Individuals with LTL values exceeding 3 standard deviations from the sample mean were excluded from the analyses, N = 3.

*Statistical Analyses*

**Observational Epidemiology**

Baseline continuous variables were compared between populations with large clone size CHIP (VAF ≧ 0.1), small clone size CHIP (VAF < 0.1), and without CHIP using ANOVA, and categorical variable associations were estimated using the chi-square test.

For the association analyses, linear regression models were used for continuous outcomes, and logistic regression models for binary outcomes.

Cox proportional hazard models were used in survival analyses with coronary artery disease as the outcome. Cox proportionality assumption was assessed by Schoenfeld. Models were adjusted for age, sex, ever smoking, body mass index (BMI), hypercholesterolemia, first 11 genetic PCs, study within TOPMed, and sequencing center (study and sequencing center are only applicable to TOPMed). Quadratic age was used as a covariate in all the models throughout this study where applicable. Study, age, and sex were stratified, and sequencing center was clustered to comply with the Cox proportionality assumption in TOPMed.

Meta analyses were performed by the fixed-effect model using "*meta*" package[57] in R. Two-sided $P < 0.05$ was considered statistically significant. Analyses were conducted using R 3.6.1 (R Foundation for Statistical Computing, Vienna, Austria).

**Mendelian randomization**

We performed bidirectional MR studies between CHIP and LTL. All procedures were consistent with the current recommendations for MR studies.[58]

Given the large scale of sample size and abundance of CHIP cases with available individual-level data, we bidirectionally performed one-sample MR in TOPMed. Instrumental variables detected in previously reported CHIP[29] and LTL[10] GWAS studies were used for the causal inference by two-stage least-squares regression. Genome-wide significance ($P < 5x10^{-8}$) was considered as the criteria for the instrumental variable (IV) assumption of robust relevance (the first assumption of Mendelian randomization) and confirmed by F statistics. IVs were pruned into independent loci < 10 Mb apart and in the linkage equilibrium ($R^2 > 0.001$ calculated in European ancestry from 1000 Genome project) using "*TwoSampleMR*" package[59] in R. Used IVs were reported in Supplementary Tables 5 and 7 for CHIP on LTL and LTL on CHIP, respectively. Age at blood draw, sex, study, sequencing center, and first eleven genetic PCs to control for population structure were included as covariates. Effect estimates for continuous exposure and outcome (LTL) were normalized to 1 standard deviation.

To mitigate the influence of reverse causality, we performed Steiger filtering to remove variants that have significantly greater association with the outcome than exposure. Some variants may be associated with exposure via first outcome and secondarily exposure. These variants should not be included as instruments for causal inference because of the violation of the exclusion-restriction assumption (the third assumption of Mendelian randomization). Previous reports showed that *TERT* locus is the leading predisposition for both CHIP and LTL, so that this locus may have significant effects on both directions. However, we did not have the knowledge about the causal directionality *a priori*. Steiger filtering calculates the variant-exposure and variant-outcome correlations and removes variants where the variant-outcome correlation is significantly greater than the variant-exposure correlation. To perform Steiger filtering, the correlations of variants with exposure and outcome were calculated. For the continuous trait (LTL), the squared correlation of each variant with LTL was calculated as the $R^2$ from the association of the trait with the variant. Cox and Snell pseudo R was calculated and squared for the binary measurement (CHIP). Steiger test was performed using the *r.test* function in R package "*psych*."[60] To apply Steiger filtering, variants with $R^2_{exposure} < R^2_{outcome}$ and $P < 0.05$ were removed.

Replication analyses

When cohorts independent from TOPMed are available for both exposure and outcome, two-sample MR with summary-level data was performed as the replication analysis. This was only applicable to the study of LTL on CHIP. GWAS summary statistics for LTL in European ancestry population[10] was used as exposure and association of outcome (CHIP) with the IVs were calculated using individual-level data in the subset of white British population from UK Biobank (N = 42,201). Study cohorts used for each MR study are summarized in Supplementary Table 8.

For two-sample MR, the analogous approach to one-sample MR with two-stage least-square regression is an inverse-variance-weighted (IVW) fixed-effects meta-analysis of the effect of each SNP on the outcome divided by the effect of this SNP on exposure, which was performed by the "*MendelianRandomization*" package[61] in R.

Sensitivity analyses

For one-sample MR, F statistics were used to support robust associations between IVs and exposure, which is the first assumption of Mendelian randomization (Relevance). The second and third assumption of Mendelian randomization requires exogeneity of the IVs, which means that IVs are not associated with outcome via horizontal pleiotropy (Exchangeability and Exclusion-restriction). Sargan test statistically tests the association of outcome with the residuals from the association of the exposure with the IVs. When the exogeneity is violated, the null hypothesis will be declined. We performed Sargan test to check the Exchangeability and Exclusion-restriction assumptions in our model. When Sargan test was violated, we tried to exclude endogenous (not exogenous) IVs by outlier exclusion. Since we do not have an appropriate method to detect outliers in one-sample MR, we excluded outliers detected in two-sample MR when using the same IVs. The model was re-evaluated after outlier exclusion.

For two-sample MR, weighted median, weighted mode, and MR-Egger were calculated using the "*MendelianRandomization*" package[61] in R. Since these methods use different assumptions to estimate the causal effect and influenced differently by biases, the stable estimates across methods indicate robust causal inference.[58] Variant effect heterogeneity assessment and outlier detection was performed via MR-PRESSO residual sum of squares (RSS) by using "*MRPRESSO*" package in R, which shows improved false-positive rates.[43] Further assessment of the reliance on a particular variant was performed by IVW-based leave-one-out analysis to detect potential outliers. All the models were re-evaluated after removing those outliers and further assessed by heterogeneity test using MR-PRESSO. Scatter plot of each variant's effect on exposure versus outcome and funnel plot to visualize the balance of horizontal pleiotropy by weak instruments were generated by "*TwoSampleMR*" package in R.[59]

Given the limited availability of GWAS study for CHIP in the European population, we could not perform conventional two-sample MR approaches for CHIP on LTL. Therefore, we performed MR-RAPS,[42] which can accommodate many weak instruments. GWAS summary statistics from the European ancestry population in TOPMed (N = 29,373)[29] was used for the exposure (CHIP), and summary statistics from ENGAGE (European Network for Genetic and Genomic Epidemiology)[41] were used for the outcome (LTL). $P < 10^{-4}$ was used for the threshold of IV discovery. R package "*TwoSampleMR*"[59] was used with the default parameters to perform MR-RAPS.

**Causal mediation analysis of LTL on CHIP associated CAD**

We performed causal mediation analysis to evaluate the proportional contribution of LTL to the association between CHIP and CAD using the R "*Mediation*" package[44] in UK Biobank (N = 44,921) and the Women's Health Initiative (WHI) (N = 3,734). For UK Biobank, the covariates used in both mediation and outcome models included age, sex, ever smoking, hypercholesterolemia, hypertension, BMI, type 2 diabetes, and first 11 genetic PCs. For WHI, covariates in both the mediation and outcome model were age at blood draw, ever smoking, race, dyslipidemia, hypertension, BMI, WHI inverse probability weight (to account for the non-random selection of women for WGS in WHI), history of hormone therapy, history of hysterectomy, and first 11 genetic PCs. Each mediation analysis model was run using 100 simulations with a quasi-Bayesian approach to estimate confidence intervals.[62]

### Acknowledgements

We gratefully acknowledge the studies and participants who provided biological samples and data for TOPMed and UK Biobank. WGS for the Trans-Omics in Precision Medicine (TOPMed) program was supported by the National Heart, Lung, and Blood Institute (NHLBI). The acknowledgements for each study in TOPMed were listed on Supplementary Table 4. Core support including centralized genomic read mapping and genotype calling, along with variant quality metrics and filtering were provided by the TOPMed Informatics Research Center (3R01HL-117626-02S1; contract HHSN268201800002I). Core support including phenotype harmonization, data management, sample-identity QC, and general program coordination were provided by the TOPMed Data Coordinating Center (R01HL-120393; U01HL-120393; contract HHSN268201800001I). Phenotype harmonization for coronary artery disease in TOPMed was supported by National Heart, Lung and Blood Institute (NHLBI) grant number R01HL146860. The views expressed in this manuscript are those of the authors and do not necessarily represent the views of the National Heart, Lung, and Blood Institute, the National Institutes of Health or the U.S. Department of Health and Human Services. All participants in the UK Biobank study provided written informed consent and Ethics approval for the UK Biobank study was obtained as described before.[63] UK Biobank Resource was used under application number 7089. Secondary use of the UK Biobank data was approved by the Massachusetts General Hospital institutional review board (protocol 2013P001840).

**NHLBI Trans-Omics for Precision Medicine (TOPMed) Consortium**

Namiko Abe[97], Francois Aguet[1], Christine Albert[98], Laura Almasy[99], Alvaro Alonso[100], Seth Ament[101], Peter Anderson[102], Pramod Anugu[103], Deborah Applebaum-Bowden[104], Kristin Ardlie[1], Dan Arking[105], Allison Ashley-Koch[106], Stella Aslibekyan[107], Paul Auer[108], Dimitrios Avramopoulos[105], Najib Ayas[109], John Barnard[110], R. Graham Barr[111], Emily Barron-Casella[105], Lucas Barwick[112], Terri Beaty[105], Gerald Beck[113], Diane Becker[24], Rebecca Beer[104], Amber Beitelshees[101], Emelia Benjamin[114], Takis Benos[115], Marcos Bezerra[116], Larry Bielak[23], Russell Bowler[117], Jennifer Brody[102], Ulrich Broeckel[118], Deborah Brown[119], Karen Bunting[97], Esteban Burchard[120], Carlos Bustamante[121], Erin Buth[17], Brian Cade[72], Jonathan Cardwell[122], Vincent Carey[123], Julie Carrier[124], Richard Casaburi[125], Juan P Casas Romero[123], James Casella[105], Peter Castaldi[88], Mark Chaffin[1], Christy Chang[101], Yi-Cheng Chang[126], Daniel Chasman[88], Bo-Juen Chen[97], Wei-Min Chen[127], Seung Hoan Choi[1], Lee-Ming Chuang[126], Mina Chung[110], Ren-Hua Chung[128], Clary Clish[1], Suzy Comhair[110], Matthew Conomos[17], Elaine Cornell[129], Carolyn Crandall[125], James Crapo[117], Jeffrey Curtis[130], Coleen Damcott[101], Sayantan Das[130], Sean David[131], Colleen Davis[102], Michelle Daya[122], Michael DeBaun[132], Scott Devine[101], Qing Duan[133], Ravi Duggirala[134], Jon Peter Durda[129], Susan Dutcher[135], Charles Eaton[136], Lynette Ekunwe[103], Adel El Boueiz[32], Serpil Erzurum[110], Charles Farber[127], Tasha Fingerlin[137], Matthew Flickinger[130], Nora Franceschini[33], Chris Frazar[102], Mao Fu[101], Stephanie M. Fullerton[102], Lucinda Fulton[135], Stacey Gabriel[1], Weiniu Gan[104], Shanshan Gao[122], Yan Gao[103], Margery Gass[92], Bruce Gelb[138], Xiaoqi (Priscilla) Geng[130], Mark Geraci[115], Soren Germer[97], Robert Gerszten[73], Auyon Ghosh[123], Richard Gibbs[139], Chris Gignoux[140], Mark Gladwin[115], David Glahn[141], Stephanie Gogarten[102], Da-Wei Gong[101], Harald Goring[19], Sharon Graw[142], Kathryn J. Gray[143], Daniel Grine[122], Yue Guan[101], Xiuqing Guo[144], Namrata Gupta[1], David Haas[145], Jeff Haessler[92], Michael Hall[103], Daniel Harris[101], Ben Heavner[17], Ryan Hernandez[120], David Herrington[146], Craig Hersh[32], Bertha Hidalgo[107], Brian Hobbs[123], John Hokanson[122], Elliott Hong[101], Karin Hoth[147], Chao (Agnes) Hsiung[128], Yi-Jen Hung[148], Haley Huston[90], Rebecca Jackson[149], Deepti Jain[102], Cashell Jaquish[104],

Min A Jhun[130], Andrew Johnson[104], Craig Johnson[102], Rich Johnston[100], Kimberly Jones[105], Hyun Min Kang[150], Sekar Kathiresan[1], Michael Kessler[101], Wonji Kim[151], Gregory L. Kinney[142], Holly Kramer[152], Christoph Lange[153], Ethan Lange[122], Leslie Lange[122], Cathy Laurie[102], Meryl LeBoff[123], Jiwon Lee[123], Seunggeun Shawn Lee[130], Wen-Jane Lee[154], Jonathon LeFaive[130], David Levine[102], Dan Levy[104], Joshua Lewis[101], Xiaohui Li[144], Yun Li[133], Henry Lin[144], Honghuang Lin[155], Keng Han Lin[130], Xihong Lin[153], Simin Liu[156], Yongmei Liu[157], Yu Liu[158], Steven Lubitz[94], Kathryn Lunetta[155], James Luo[104], Ulysses Magalang[159], Michael Mahaney[19], Barry Make[105], Ani Manichaikul[127], Lauren Margolin[1], Lisa Martin[160], Susan Mathai[122], Susanne May[17], Patrick McArdle[101], Merry-Lynn McDonald[107], Sean McFarland[151], Daniel McGoldrick[102], Caitlin McHugh[17], Becky McNeil[161], Hao Mei[103], Luisa Mestroni[142], Emmanuel Mignot[162], Julie Mikulla[104], Nancy Min[103], Mollie Minear[104], Matt Moll[88], May E. Montasser[101], Courtney Montgomery[163], Arden Moscati[138], Stanford Mwasongwe[103], Josyf C Mychaleckyj[127], Girish Nadkarni[138], Rakhi Naik[105], Sergei Nekhai[164], Sarah C. Nelson[17], Bonnie Neltner[122], Deborah Nickerson[102], Jeff O'Connell[101], Tim O'Connor[101], Heather Ochs-Balcom[165], Allan Pack[166], David T. Paik[158], James Pankow[167], George Papanicolaou[104], Cora Parker[168], Afshin Parsa[101], Marco Perez[140], James Perry[101], Ulrike Peters[92], Lawrence S Phillips[100], Toni Pollin[101], Julia Powers Becker[122], Pankaj Qasba[104], Zhaohui Qin[100], Nicholas Rafaels[122], D.C. Rao[135], Laura Rasmussen-Torvik[169], Aakrosh Ratan[127], Robert Reed[101], Elizabeth Regan[117], Ken Rice[102], Carolina Roselli[1], Ingo Ruczinski[105], Pamela Russell[122], Sarah Ruuska[90], Kathleen Ryan[101], Ester Cerdeira Sabino[170], Danish Saleheen[111], Shabnam Salimi[101], Steven Salzberg[105], Kevin Sandow[144], Vijay G. Sankaran[171], Christopher Scheller[130], Ellen Schmidt[130], Karen Schwander[135], David Schwartz[122], Frank Sciurba[115], Christine Seidman[172], Jonathan Seidman[172], Vivien Sheehan[173], Stephanie L. Sherman[174], Amol Shetty[101], Aniket Shetty[122], Wayne Hui-Heng Sheu[154], Brian Silver[175], Josh Smith[102], Tanja Smith[97], Sylvia Smoller[82], Beverly Snively[176], Michael Snyder[140], Tamar Sofer[123], Nona Sotoodehnia[102], Garrett Storm[122], Elizabeth Streeten[101], Jessica Lasky Su[123], Yun Ju Sung[135], Jody Sylvia[123], Adam Szpiro[102], Carole Sztalryd[101], Daniel Taliun[130], Hua Tang[140], Kent D. Taylor[81],

Matthew Taylor[142], Simeon Taylor[101], Marilyn Telen[106], Timothy A. Thornton[102], Machiko Threlkeld[177], Lesley Tinker[92], David Tirschwell[102], Sarah Tishkoff[178], Catherine Tong[17], Michael Tsai[167], Dhananjay Vaidya[105], David Van Den Berg[179], Peter VandeHaar[130], Scott Vrieze[167], Tarik Walker[122], Robert Wallace[147], Avram Walts[122], Fei Fei Wang[102], Heming Wang[123], Karol Watson[125], Bruce Weir[102], Lu-Chen Weng[2], Jennifer Wessel[180], Cristen Willer[181], Kayleen Williams[17], L. Keoki Williams[182], Carla Wilson[123], James Wilson[183], Joseph Wu[158], Huichun Xu[101], Ivana Yang[122], Rongze Yang[101], Norann Zaghloul[101], Yingze Zhang[184], Snow Xueyan Zhao[117], Degui Zhi[185], Xiang Zhou[130], Xiaofeng Zhu[186], Michael Zody[97] & Sebastian Zoellner[11]

[97]New York Genome Center, New York, NY, USA. [98]Cedars Sinai, Boston, MA, USA. [99]Children's Hospital of Philadelphia, University of Pennsylvania, Philadelphia, PA, USA. [100]Emory University, Atlanta, GA, USA. [101]University of Maryland, Baltimore, MD, USA. [102]University of Washington, Seattle, WA, USA. [103]University of Mississippi, Jackson, MS, USA. [104]National Heart, Lung, and Blood Institute, National Institutes of Health, Bethesda, MD, USA. [105]Johns Hopkins University, Baltimore, MD, USA. [106]Duke University, Durham, NC, USA. [107]University of Alabama, Birmingham, AL, USA. [108]University of Wisconsin Milwaukee, Milwaukee, WI, USA. [109]Department of Medicine, Providence Health Care, Vancouver, BC, Canada. [110]Cleveland Clinic, Cleveland, OH, USA. [111]Columbia University, New York, NY, USA. [112]LTRC, The Emmes Corporation, Rockville, MD, USA. [113]Department of Quantitative Health Sciences, Cleveland Clinic, Cleveland, OH, USA. [114]Departments of Medicine and Epidemiology, Boston University School of Medicine, Boston, MA, USA. [115]University of Pittsburgh, Pittsburgh, PA, USA. [116]Fundação de Hematologia e Hemoterapia de Pernambuco (Hemope), Recife, Brazil. [117]National Jewish Health, Denver, CO, USA. [118]Medical College of Wisconsin, Milwaukee, WI, USA. [119]Department of Pediatrics, University of Texas Health at Houston, Houston, TX, USA. [120]University of California, San Francisco, San Francisco, CA, USA. [121]Department of Biomedical Data Science, Stanford University, Stanford, CA, USA. [122]University of

Colorado at Denver, Denver, CO, USA. [123]Brigham and Women's Hospital, Boston, MA, USA. [124]University of Montreal, Montreal, QC, Canada. [125]University of California, Los Angeles, Los Angeles, CA, USA. [126]National Taiwan University Hospital, National Taiwan University, Taipei, Taiwan. [127]University of Virginia, Charlottesville, VA, USA. [128]National Health Research Institute Taiwan, Miaoli County, Taiwan. [129]University of Vermont, Burlington, VT, USA. [130]University of Michigan, Ann Arbor, MI, USA. [131]University of Chicago, Chicago, IL, USA. [132]Vanderbilt University, Nashville, TN, USA. [133]University of North Carolina, Chapel Hill, NC, USA. [134]University of Texas Rio Grande Valley School of Medicine, Edinburg, TX, USA. [135]Washington University in St Louis, St Louis, MO, USA. [136]Brown University, Providence, RI, USA. [137]Center for Genes, Environment and Health, National Jewish Health, Denver, CO, USA. [138]Icahn School of Medicine at Mount Sinai, New York, NY, USA. [139]Baylor College of Medicine Human Genome Sequencing Center, Houston, TX, USA. [140]Stanford University, Stanford, CA, USA. [141]Department of Psychiatry, Boston Children's Hospital, Harvard Medical School, Boston, MA, USA. [142]University of Colorado Anschutz Medical Campus, Aurora, CO, USA. [143]Department of Obstetrics and Gynecology, Brigham and Women's Hospital, Boston, MA, USA. [144]Lundquist Institute, Torrance, CA, USA. [145]Department of Obstetrics & Gynecology, Indiana University, Indianapolis, IN, USA. [146]Wake Forest Baptist Health, Winston-Salem, NC, USA. [147]University of Iowa, Iowa City, IA, USA. [148]Tri-Service General Hospital National Defense Medical Center, Taipei, Taiwan. [149]Division of Endocrinology, Diabetes and Metabolism, Department of Internal Medicine, Oklahoma State University Medical Center, Columbus, OH, USA. [150]Department of Biostatistics, University of Michigan, Ann Arbor, MI, USA. [151]Harvard University, Cambridge, MA, USA. [152]Department of Public Health Sciences, Loyola University, Maywood, IL, USA. [153]Harvard T.H. Chan School of Public Health, Boston, MA, USA. [154]Taichung Veterans General Hospital Taiwan, Taichung City, Taiwan. [155]Boston University, Boston, MA, USA. [156]Department of Epidemiology and Medicine, Brown University, Providence, RI, USA. [157]Division of Cardiology, Department of Medicine, Duke

University, Durham, NC, USA. [158]Cardiovascular Institute, Stanford University, Stanford, CA, USA. [159]Division of Pulmonary, Critical Care and Sleep Medicine, Oklahoma State University Medical Center, Columbus, OH, USA. [160]George Washington University, Washington, USA. [161]RTI International, NC, USA. [162]Center For Sleep Sciences and Medicine, Stanford University, Palo Alto, CA, USA. [163]Department of Genes and Human Disease, Oklahoma Medical Research Foundation, Oklahoma City, OK, USA. [164]Howard University, Washington, USA. [165]University at Buffalo, Buffalo, NY, USA. [166]Division of Sleep Medicine, Department of Medicine, University of Pennsylvania, Philadelphia, PA, USA. [167]University of Minnesota, Minneapolis, MN, USA. [168]Biostatistics and Epidemiology Division, RTI International, Research Triangle Park, NC, USA. [169]Northwestern University, Chicago, IL, USA. [170]Faculdade de Medicina, Universidade de Sao Paulo, Sao Paulo, Brazil. [171]Division of Hematology/Oncology, Boston Children's Hospital and Department of Pediatric Oncology, Dana-Farber Cancer Institute, Boston, MA, USA. [172]Harvard Medical School, Boston, MA, USA. [173]Department of Pediatrics, Baylor College of Medicine, Atlanta, GA, USA. [174]Department of Human Genetics, Emory University, Atlanta, GA, USA. [175]UMass Memorial Medical Center, Worcester, MA, USA. [176]Department of Biostatistics and Data Science, Wake Forest Baptist Health, Winston-Salem, NC, USA. [177]Department of Genome Sciences, University of Washington, Seattle, WA, USA. [178]Department of Genetics, University of Pennsylvania, Philadelphia, PA, USA. [179]USC Methylation Characterization Center, University of Southern California, Los Angeles, CA, USA. [180]Department of Epidemiology, Indiana University, Indianapolis, IN, USA. [181]Department of Internal Medicine, University of Michigan, Ann Arbor, MI, USA. [182]Henry Ford Health System, Detroit, MI, USA. [183]Department of Cardiology, Beth Israel Deaconess Medical Center, Cambridge, MA, USA. [184]Department of Medicine, University of Pittsburgh, Pittsburgh, PA, USA. [185]University of Texas Health at Houston, Houston, TX, USA. [186]Department of Population and Quantitative Health Sciences, Case Western Reserve University, Cleveland, OH, USA.

**NHLBI Trans-Omics for Precision Medicine (TOPMed) Consortium**

Namiko Abe[97], Francois Aguet[1], Christine Albert[98], Laura Almasy[99], Alvaro Alonso[100], Seth Ament[101], Peter Anderson[102], Pramod Anugu[103], Deborah Applebaum-Bowden[104], Kristin Ardlie[1], Dan Arking[105], Allison Ashley-Koch[106], Stella Aslibekyan[107], Paul Auer[108], Dimitrios Avramopoulos[105], Najib Ayas[109], John Barnard[110], R. Graham Barr[111], Emily Barron-Casella[105], Lucas Barwick[112], Terri Beaty[105], Gerald Beck[113], Diane Becker[24], Rebecca Beer[104], Amber Beitelshees[101], Emelia Benjamin[114], Takis Benos[115], Marcos Bezerra[116], Larry Bielak[23], Russell Bowler[117], Jennifer Brody[102], Ulrich Broeckel[118], Deborah Brown[119], Karen Bunting[97], Esteban Burchard[120], Carlos Bustamante[121], Erin Buth[17], Brian Cade[72], Jonathan Cardwell[122], Vincent Carey[123], Julie Carrier[124], Richard Casaburi[125], Juan P Casas Romero[123], James Casella[105], Peter Castaldi[88], Mark Chaffin[1], Christy Chang[101], Yi-Cheng Chang[126], Daniel Chasman[88], Bo-Juen Chen[97], Wei-Min Chen[127], Seung Hoan Choi[1], Lee-Ming Chuang[126], Mina Chung[110], Ren-Hua Chung[128], Clary Clish[1], Suzy Comhair[110], Matthew Conomos[17], Elaine Cornell[129], Carolyn Crandall[125], James Crapo[117], Jeffrey Curtis[130], Coleen Damcott[101], Sayantan Das[130], Sean David[131], Colleen Davis[102], Michelle Daya[122], Michael DeBaun[132], Scott Devine[101], Qing Duan[133], Ravi Duggirala[134], Jon Peter Durda[129], Susan Dutcher[135], Charles Eaton[136], Lynette Ekunwe[103], Adel El Boueiz[32], Serpil Erzurum[110], Charles Farber[127], Tasha Fingerlin[137], Matthew Flickinger[130], Nora Franceschini[33], Chris Frazar[102], Mao Fu[101], Stephanie M. Fullerton[102], Lucinda Fulton[135], Stacey Gabriel[1], Weiniu Gan[104], Shanshan Gao[122], Yan Gao[103], Margery Gass[92], Bruce Gelb[138], Xiaoqi (Priscilla) Geng[130], Mark Geraci[115], Soren Germer[97], Robert Gerszten[73], Auyon Ghosh[123], Richard Gibbs[139], Chris Gignoux[140], Mark Gladwin[115], David Glahn[141], Stephanie Gogarten[102], Da-Wei Gong[101], Harald Goring[19], Sharon Graw[142], Kathryn J. Gray[143], Daniel Grine[122], Yue Guan[101], Xiuqing Guo[144], Namrata Gupta[1], David Haas[145], Jeff Haessler[92], Michael Hall[103], Daniel Harris[101], Ben Heavner[17], Ryan Hernandez[120], David Herrington[146], Craig Hersh[32], Bertha Hidalgo[107], Brian Hobbs[123], John Hokanson[122], Elliott Hong[101], Karin Hoth[147], Chao (Agnes) Hsiung[128], Yi-Jen Hung[148], Haley Huston[90], Rebecca Jackson[149], Deepti

Jain[102], Cashell Jaquish[104], Min A Jhun[130], Andrew Johnson[104], Craig Johnson[102], Rich Johnston[100], Kimberly Jones[105], Hyun Min Kang[150], Sekar Kathiresan[1], Michael Kessler[101], Wonji Kim[151], Gregory L. Kinney[142], Holly Kramer[152], Christoph Lange[153], Ethan Lange[122], Leslie Lange[122], Cathy Laurie[102], Meryl LeBoff[123], Jiwon Lee[123], Seunggeun Shawn Lee[130], Wen-Jane Lee[154], Jonathon LeFaive[130], David Levine[102], Dan Levy[104], Joshua Lewis[101], Xiaohui Li[144], Yun Li[133], Henry Lin[144], Honghuang Lin[155], Keng Han Lin[130], Xihong Lin[153], Simin Liu[156], Yongmei Liu[157], Yu Liu[158], Steven Lubitz[94], Kathryn Lunetta[155], James Luo[104], Ulysses Magalang[159], Michael Mahaney[19], Barry Make[105], Ani Manichaikul[127], Lauren Margolin[1], Lisa Martin[160], Susan Mathai[122], Susanne May[17], Patrick McArdle[101], Merry-Lynn McDonald[107], Sean McFarland[151], Daniel McGoldrick[102], Caitlin McHugh[17], Becky McNeil[161], Hao Mei[103], Luisa Mestroni[142], Emmanuel Mignot[162], Julie Mikulla[104], Nancy Min[103], Mollie Minear[104], Matt Moll[88], May E. Montasser[101], Courtney Montgomery[163], Arden Moscati[138], Stanford Mwasongwe[103], Josyf C Mychaleckyj[127], Girish Nadkarni[138], Rakhi Naik[105], Sergei Nekhai[164], Sarah C. Nelson[17], Bonnie Neltner[122], Deborah Nickerson[102], Jeff O'Connell[101], Tim O'Connor[101], Heather Ochs-Balcom[165], Allan Pack[166], David T. Paik[158], James Pankow[167], George Papanicolaou[104], Cora Parker[168], Afshin Parsa[101], Marco Perez[140], James Perry[101], Ulrike Peters[92], Lawrence S Phillips[100], Toni Pollin[101], Julia Powers Becker[122], Pankaj Qasba[104], Zhaohui Qin[100], Nicholas Rafaels[122], D.C. Rao[135], Laura Rasmussen-Torvik[169], Aakrosh Ratan[127], Robert Reed[101], Elizabeth Regan[117], Ken Rice[102], Carolina Roselli[1], Ingo Ruczinski[105], Pamela Russell[122], Sarah Ruuska[90], Kathleen Ryan[101], Ester Cerdeira Sabino[170], Danish Saleheen[111], Shabnam Salimi[101], Steven Salzberg[105], Kevin Sandow[144], Vijay G. Sankaran[171], Christopher Scheller[130], Ellen Schmidt[130], Karen Schwander[135], David Schwartz[122], Frank Sciurba[115], Christine Seidman[172], Jonathan Seidman[172], Vivien Sheehan[173], Stephanie L. Sherman[174], Amol Shetty[101], Aniket Shetty[122], Wayne Hui-Heng Sheu[154], Brian Silver[175], Josh Smith[102], Tanja Smith[97], Sylvia Smoller[82], Beverly Snively[176], Michael Snyder[140], Tamar Sofer[123], Nona Sotoodehnia[102], Garrett Storm[122], Elizabeth Streeten[101], Jessica Lasky Su[123], Yun Ju Sung[135], Jody Sylvia[123], Adam Szpiro[102], Carole Sztalryd[101],

Daniel Taliun[130], Hua Tang[140], Kent D. Taylor[81], Matthew Taylor[142], Simeon Taylor[101], Marilyn Telen[106], Timothy A. Thornton[102], Machiko Threlkeld[177], Lesley Tinker[92], David Tirschwell[102], Sarah Tishkoff[178], Catherine Tong[17], Michael Tsai[167], Dhananjay Vaidya[105], David Van Den Berg[179], Peter VandeHaar[130], Scott Vrieze[167], Tarik Walker[122], Robert Wallace[147], Avram Walts[122], Fei Fei Wang[102], Heming Wang[123], Karol Watson[125], Bruce Weir[102], Lu-Chen Weng[2], Jennifer Wessel[180], Cristen Willer[181], Kayleen Williams[17], L. Keoki Williams[182], Carla Wilson[123], James Wilson[183], Joseph Wu[158], Huichun Xu[101], Ivana Yang[122], Rongze Yang[101], Norann Zaghloul[101], Yingze Zhang[184], Snow Xueyan Zhao[117], Degui Zhi[185], Xiang Zhou[130], Xiaofeng Zhu[186], Michael Zody[97] & Sebastian Zoellner[11]

[97]New York Genome Center, New York, NY, USA. [98]Cedars Sinai, Boston, MA, USA. [99]Children's Hospital of Philadelphia, University of Pennsylvania, Philadelphia, PA, USA. [100]Emory University, Atlanta, GA, USA. [101]University of Maryland, Baltimore, MD, USA. [102]University of Washington, Seattle, WA, USA. [103]University of Mississippi, Jackson, MS, USA. [104]National Heart, Lung, and Blood Institute, National Institutes of Health, Bethesda, MD, USA. [105]Johns Hopkins University, Baltimore, MD, USA. [106]Duke University, Durham, NC, USA. [107]University of Alabama, Birmingham, AL, USA. [108]University of Wisconsin Milwaukee, Milwaukee, WI, USA. [109]Department of Medicine, Providence Health Care, Vancouver, BC, Canada. [110]Cleveland Clinic, Cleveland, OH, USA. [111]Columbia University, New York, NY, USA. [112]LTRC, The Emmes Corporation, Rockville, MD, USA. [113]Department of Quantitative Health Sciences, Cleveland Clinic, Cleveland, OH, USA. [114]Departments of Medicine and Epidemiology, Boston University School of Medicine, Boston, MA, USA. [115]University of Pittsburgh, Pittsburgh, PA, USA. [116]Fundação de Hematologia e Hemoterapia de Pernambuco (Hemope), Recife, Brazil. [117]National Jewish Health, Denver, CO, USA. [118]Medical College of Wisconsin, Milwaukee, WI, USA. [119]Department of Pediatrics, University of Texas Health at Houston, Houston, TX, USA. [120]University of California,

San Francisco, San Francisco, CA, USA. [121]Department of Biomedical Data Science, Stanford University, Stanford, CA, USA. [122]University of Colorado at Denver, Denver, CO, USA. [123]Brigham and Women's Hospital, Boston, MA, USA. [124]University of Montreal, Montreal, QC, Canada. [125]University of California, Los Angeles, Los Angeles, CA, USA. [126]National Taiwan University Hospital, National Taiwan University, Taipei, Taiwan. [127]University of Virginia, Charlottesville, VA, USA. [128]National Health Research Institute Taiwan, Miaoli County, Taiwan. [129]University of Vermont, Burlington, VT, USA. [130]University of Michigan, Ann Arbor, MI, USA. [131]University of Chicago, Chicago, IL, USA. [132]Vanderbilt University, Nashville, TN, USA. [133]University of North Carolina, Chapel Hill, NC, USA. [134]University of Texas Rio Grande Valley School of Medicine, Edinburg, TX, USA. [135]Washington University in St Louis, St Louis, MO, USA. [136]Brown University, Providence, RI, USA. [137]Center for Genes, Environment and Health, National Jewish Health, Denver, CO, USA. [138]Icahn School of Medicine at Mount Sinai, New York, NY, USA. [139]Baylor College of Medicine Human Genome Sequencing Center, Houston, TX, USA. [140]Stanford University, Stanford, CA, USA. [141]Department of Psychiatry, Boston Children's Hospital, Harvard Medical School, Boston, MA, USA. [142]University of Colorado Anschutz Medical Campus, Aurora, CO, USA. [143]Department of Obstetrics and Gynecology, Brigham and Women's Hospital, Boston, MA, USA. [144]Lundquist Institute, Torrance, CA, USA. [145]Department of Obstetrics & Gynecology, Indiana University, Indianapolis, IN, USA. [146]Wake Forest Baptist Health, Winston-Salem, NC, USA. [147]University of Iowa, Iowa City, IA, USA. [148]Tri-Service General Hospital National Defense Medical Center, Taipei, Taiwan. [149]Division of Endocrinology, Diabetes and Metabolism, Department of Internal Medicine, Oklahoma State University Medical Center, Columbus, OH, USA. [150]Department of Biostatistics, University of Michigan, Ann Arbor, MI, USA. [151]Harvard University, Cambridge, MA, USA. [152]Department of Public Health Sciences, Loyola University, Maywood, IL, USA. [153]Harvard T.H. Chan School of Public Health, Boston, MA, USA. [154]Taichung Veterans General Hospital Taiwan, Taichung City, Taiwan. [155]Boston University, Boston, MA, USA.

[156]Department of Epidemiology and Medicine, Brown University, Providence, RI, USA. [157]Division of Cardiology, Department of Medicine, Duke University, Durham, NC, USA. [158]Cardiovascular Institute, Stanford University, Stanford, CA, USA. [159]Division of Pulmonary, Critical Care and Sleep Medicine, Oklahoma State University Medical Center, Columbus, OH, USA. [160]George Washington University, Washington, USA. [161]RTI International, NC, USA. [162]Center For Sleep Sciences and Medicine, Stanford University, Palo Alto, CA, USA. [163]Department of Genes and Human Disease, Oklahoma Medical Research Foundation, Oklahoma City, OK, USA. [164]Howard University, Washington, USA. [165]University at Buffalo, Buffalo, NY, USA. [166]Division of Sleep Medicine, Department of Medicine, University of Pennsylvania, Philadelphia, PA, USA. [167]University of Minnesota, Minneapolis, MN, USA. [168]Biostatistics and Epidemiology Division, RTI International, Research Triangle Park, NC, USA. [169]Northwestern University, Chicago, IL, USA. [170]Faculdade de Medicina, Universidade de Sao Paulo, Sao Paulo, Brazil. [171]Division of Hematology/Oncology, Boston Children's Hospital and Department of Pediatric Oncology, Dana-Farber Cancer Institute, Boston, MA, USA. [172]Harvard Medical School, Boston, MA, USA. [173]Department of Pediatrics, Baylor College of Medicine, Atlanta, GA, USA. [174]Department of Human Genetics, Emory University, Atlanta, GA, USA. [175]UMass Memorial Medical Center, Worcester, MA, USA. [176]Department of Biostatistics and Data Science, Wake Forest Baptist Health, Winston-Salem, NC, USA. [177]Department of Genome Sciences, University of Washington, Seattle, WA, USA. [178]Department of Genetics, University of Pennsylvania, Philadelphia, PA, USA. [179]USC Methylation Characterization Center, University of Southern California, Los Angeles, CA, USA. [180]Department of Epidemiology, Indiana University, Indianapolis, IN, USA. [181]Department of Internal Medicine, University of Michigan, Ann Arbor, MI, USA. [182]Henry Ford Health System, Detroit, MI, USA. [183]Department of Cardiology, Beth Israel Deaconess Medical Center, Cambridge, MA, USA. [184]Department of Medicine, University of Pittsburgh, Pittsburgh, PA, USA. [185]University of Texas

Health at Houston, Houston, TX, USA. [186]Department of Population and Quantitative Health Sciences, Case Western Reserve University, Cleveland, OH, USA.

**Supplementary Fig. 1: Generation of the study population for observational studies.**

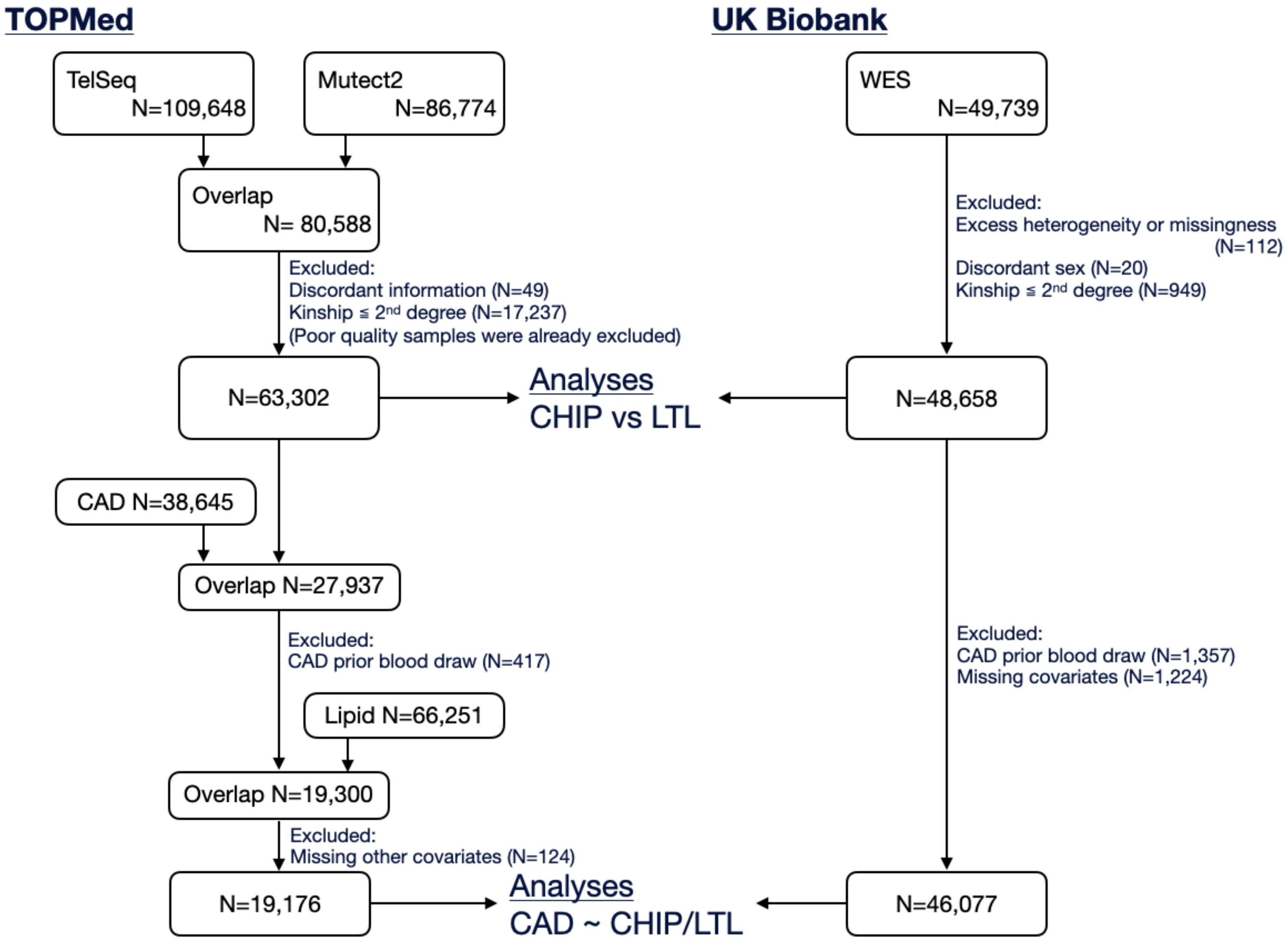


The flow chart for generating the study population used for observational studies in TOPMed and UK Biobank. CAD: coronary artery disease, CHIP: clonal hematopoiesis of intermediate potential, LTL: leukocyte telomere length, TOPMed: Trans-Omics for Precision Medicine, WES: whole-exome sequencing.

**Supplementary Fig. 2: Batch correction of estimated leukocyte telomere length from whole-exome sequencing.**

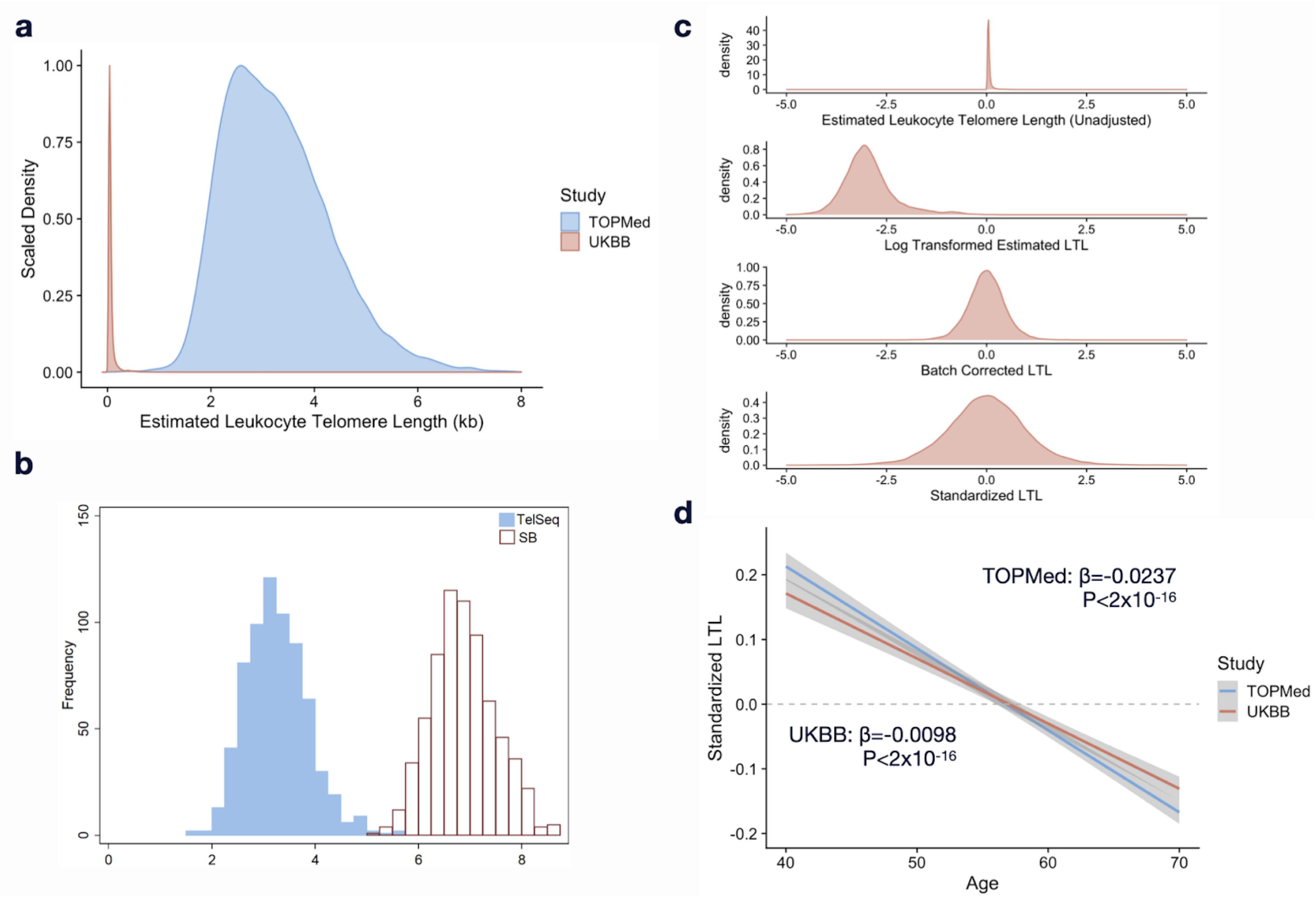


**a** Telomere length was estimated by TelSeq using whole-exome sequence data in UK Biobank (N=48,658) is shorter than TOPMed (N=63,302) using whole genome sequencing. **b** Estimated LTL by TelSeq using WGS is shorter than the measurement by southern blot when directly compared in the same individuals in a subset of WHI cohort (N=686) **c** The estimated LTL was log transformed, regressed out by first 9 PCs, and standardized for mean = 0 and standard deviation =1 for each cohort. **d** Correlation between LTL and age in TOPMed and UK Biobank. A subset of age 40 to 70 years were compared and displayed. Estimates are adjusted by age, sex, ever smoking, body mass index, study cohort, and sequencing center (cohort and sequencing center are only applicable to TOPMed). kb: kilo base, LTL: leukocyte telomere length, SB: southern blot, TOPMed: Trans-Omics for Precision Medicine, UKBB: UK Biobank, WHI: Women's Health Initiative.

**Supplementary Fig. 3: SNPs' effect sizes for associations with LTL are strongly correlated between direct measurement and WES-derived estimation.**

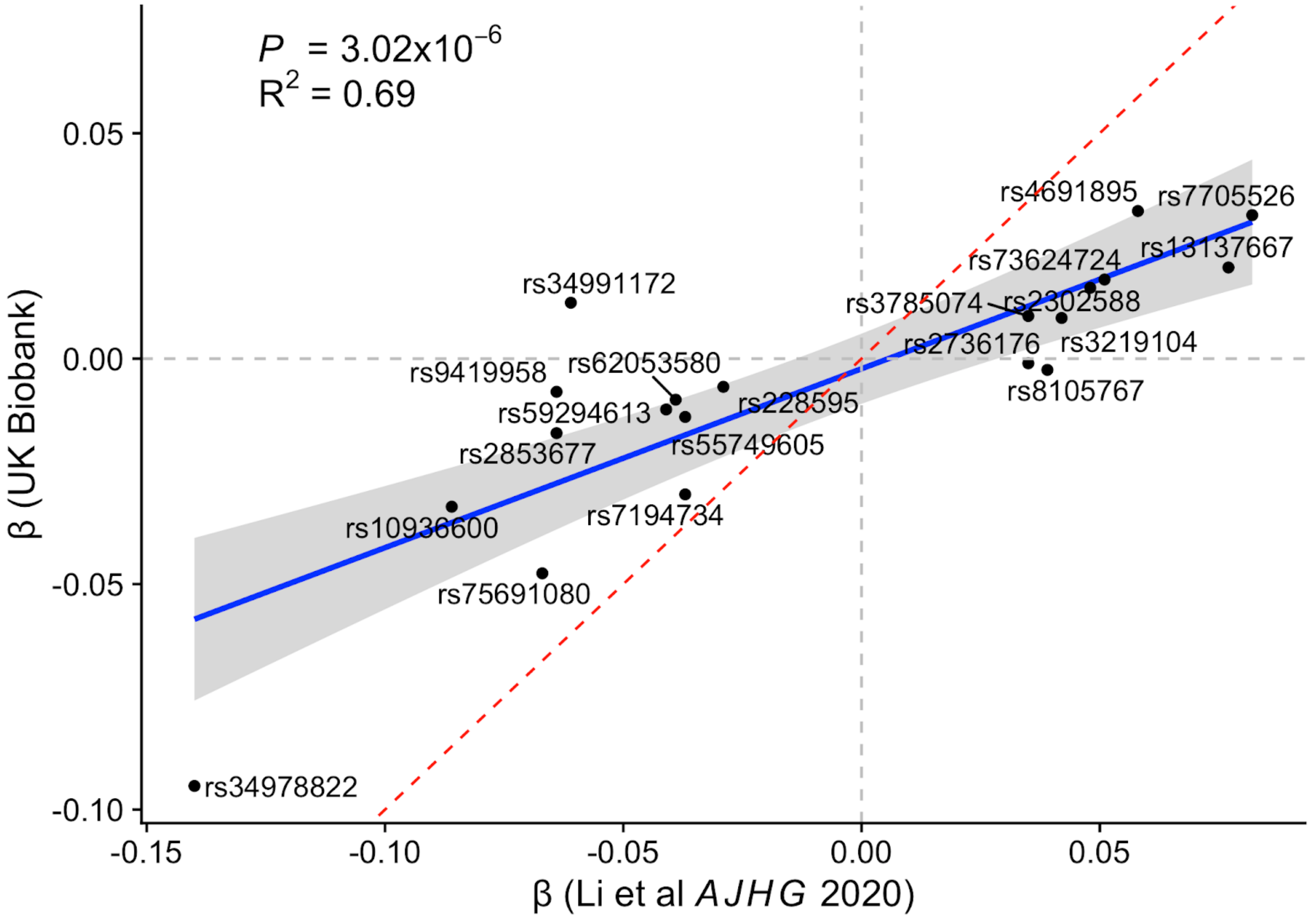


Correlation of estimates in the effect of variants for LTL between our data from UK Biobank and previous report (Li et al *AJHG* 2020). Effect sizes were calculated in a subset of white British population in UK Biobank (N=42,201) and adjusted by age, sex, and first 11 genetic principal components. Red dotted lines represent equality between both data (x=y). LTL: leukocyte telomere length, SNP: single nucleotide polymorphism, UKBB: UK Biobank, WES: whole-exome sequencing.

**Supplementary Fig. 4: Longer LTL is associated with reduced CHIP prevalence, VAF, and incident CAD risk.**

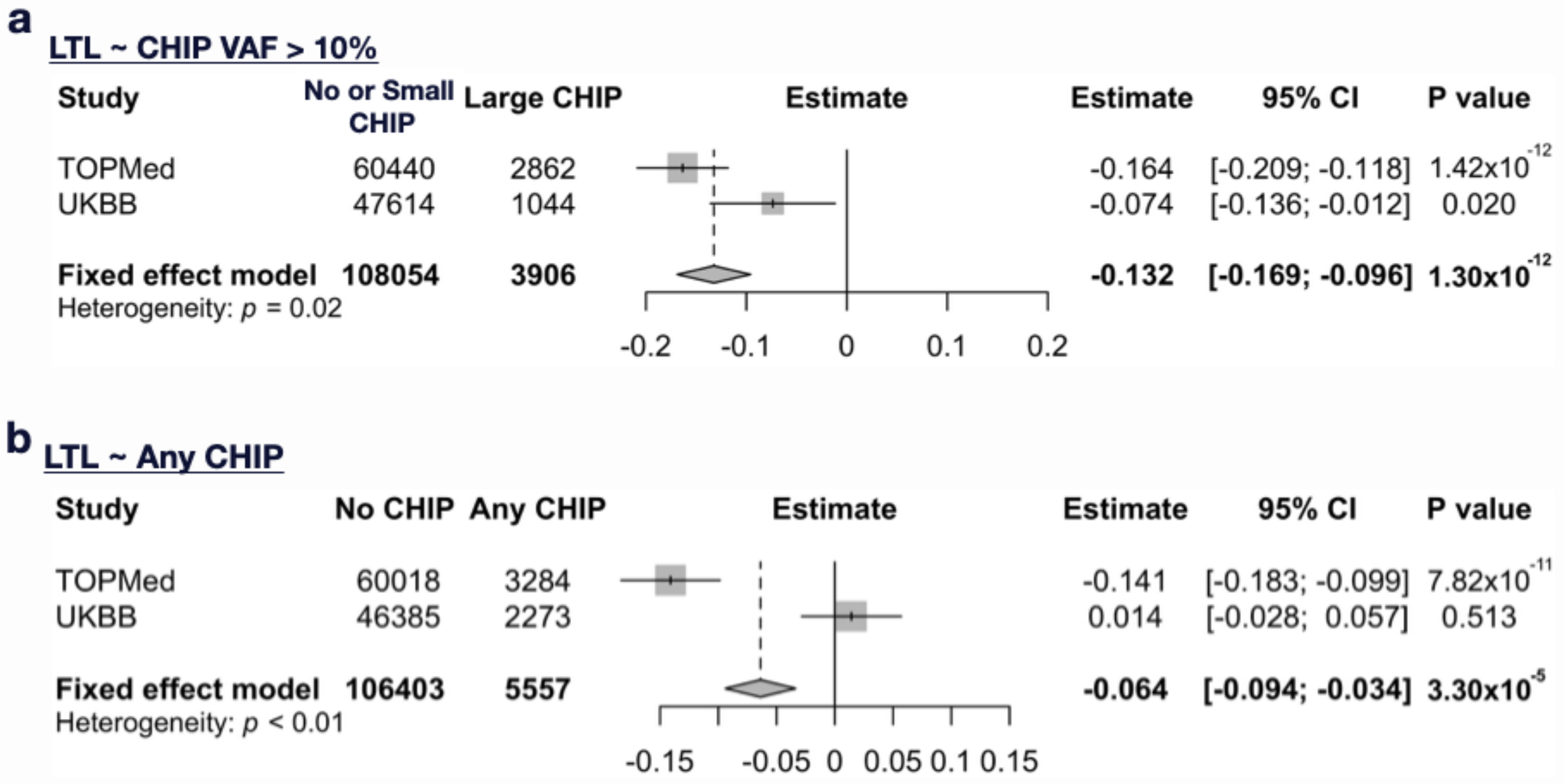


**a,b** For the outcomes **a** CHIP with VAF > 0.1 and **b** any CHIP, the associations with LTL were assessed by linear regression model both in TOPMed and UK Biobank, then meta-analyzed. Both models were adjusted with age, sex, ever smoking, body mass index, first 11 PCs, study, and sequencing center (study and sequencing center are only applicable to TOPMed). CHIP: clonal hematopoiesis of indeterminate potential, CI: confidence interval, HR: hazard ratio, LTL: leukocyte telomere length, PC: principal component, TOPMed: Trans-Omics for Precision Medicine, UKBB: UK Biobank, VAF: variant allele frequency.

**Supplementary Fig. 5: Effect of CHIP (VAF>10%) on LTL per CHIP gene (most frequently mutated 10 genes)**

| Gene | Study | No CHIP | CHIP (VAF>10%) | Estimate | 95% CI | P value |
|---|---|---|---|---|---|---|
| DNMT3A | TOPMed | 60018 | 1496 | -0.0189 | [ -0.0798 : 0.0421 ] | 0.544 |
| | UKBB | 46385 | 572 | -0.0589 | [ -0.142 : 0.0243 ] | 0.165 |
| | **Fixed effect model** | **106403** | **2068** | **-0.0328** | **[ -0.082 : 0.0163 ]** | **0.19** |
| TET2 | TOPMed | 60018 | 510 | -0.285 | [ -0.386 : -0.183 ] | 4.33e-08 |
| | UKBB | 46385 | 221 | -0.0544 | [ -0.189 : 0.0802 ] | 0.428 |
| | **Fixed effect model** | **106403** | **731** | **-0.201** | **[ -0.282 : -0.12 ]** | **1.25e-06** |
| ASXL1 | TOPMed | 60018 | 233 | -0.426 | [ -0.572 : -0.279 ] | 1.15e-08 |
| | UKBB | 46385 | 84 | -0.0667 | [ -0.284 : 0.151 ] | 0.547 |
| | **Fixed effect model** | **106403** | **317** | **-0.314** | **[ -0.435 : -0.193 ]** | **3.91e-07** |
| PPM1D | TOPMed | 60018 | 104 | -0.613 | [ -0.852 : -0.373 ] | 5.53e-07 |
| | UKBB | 46385 | 21 | -0.147 | [ -0.574 : 0.279 ] | 0.499 |
| | **Fixed effect model** | **106403** | **125** | **-0.501** | **[ -0.71 : -0.292 ]** | **2.64e-06** |
| JAK2 | TOPMed | 60018 | 92 | -0.798 | [ -1.05 : -0.543 ] | 8.93e-10 |
| | UKBB | 46385 | 9 | -0.197 | [ -0.848 : 0.455 ] | 0.554 |
| | **Fixed effect model** | **106403** | **101** | **-0.718** | **[ -0.955 : -0.48 ]** | **3.17e-09** |
| SF3B1 | TOPMed | 60018 | 72 | 0.00365 | [ -0.261 : 0.268 ] | 0.978 |
| | UKBB | 46385 | 14 | -0.485 | [ -1.01 : 0.0374 ] | 0.0688 |
| | **Fixed effect model** | **106403** | **86** | **-0.0959** | **[ -0.332 : 0.14 ]** | **0.425** |
| TP53 | TOPMed | 60018 | 54 | -0.665 | [ -1 : -0.326 ] | 0.000121 |
| | UKBB | 46385 | 21 | -0.395 | [ -0.832 : 0.042 ] | 0.0765 |
| | **Fixed effect model** | **106403** | **75** | **-0.563** | **[ -0.831 : -0.296 ]** | **3.72e-05** |
| SRSF2 | TOPMed | 60018 | 50 | -0.0349 | [ -0.36 : 0.29 ] | 0.833 |
| | UKBB | 46385 | 8 | -0.544 | [ -1.23 : 0.147 ] | 0.123 |
| | **Fixed effect model** | **106403** | **58** | **-0.127** | **[ -0.421 : 0.167 ]** | **0.397** |
| GNB1 | TOPMed | 60018 | 26 | -0.281 | [ -0.76 : 0.198 ] | 0.25 |
| | UKBB | 46385 | 11 | -0.302 | [ -0.891 : 0.287 ] | 0.314 |
| | **Fixed effect model** | **106403** | **37** | **-0.29** | **[ -0.661 : 0.0819 ]** | **0.127** |
| GNAS | TOPMed | 60018 | 22 | 0.0755 | [ -0.472 : 0.623 ] | 0.787 |
| | UKBB | 46385 | 6 | 0.0677 | [ -0.73 : 0.865 ] | 0.868 |
| | **Fixed effect model** | **106403** | **28** | **0.073** | **[ -0.378 : 0.524 ]** | **0.751** |

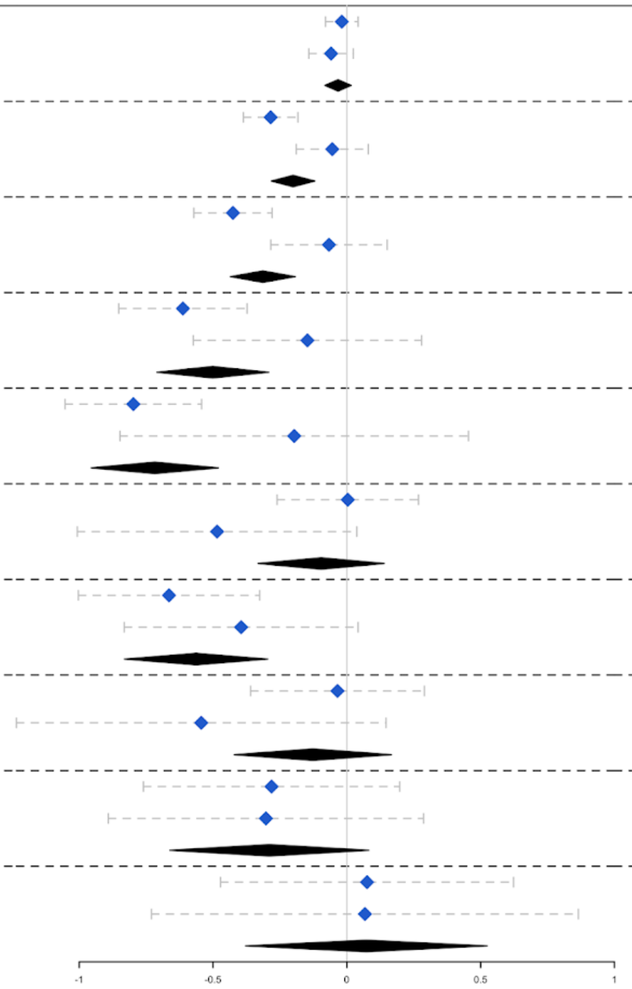


Effect estimate of large clone size CHIP (VAF > 10 %) on LTL was assessed in each CHIP gene. Linear regression model was adjusted with age, sex, smoking, BMI, first 11 genetic principal components, study, and sequencing center in both cohort and meta-analyzed using fixed effect model (study and sequencing center are only applicable to TOPMed). The 10 most frequently mutated genes are displayed. Data for the other CHIP genes is reported in Supplementary Table 2. CHIP: clonal hematopoiesis of indeterminate potential, CI: confidence interval, LTL: leukocyte telomere length, TOPMed: Trans-Omics for Precision Medicine, UKBB: UK Biobank, VAF: variant allele frequency.

**Supplementary Fig. 6: Association of CHIP with LTL per number of genes affected by CHIP mutations**

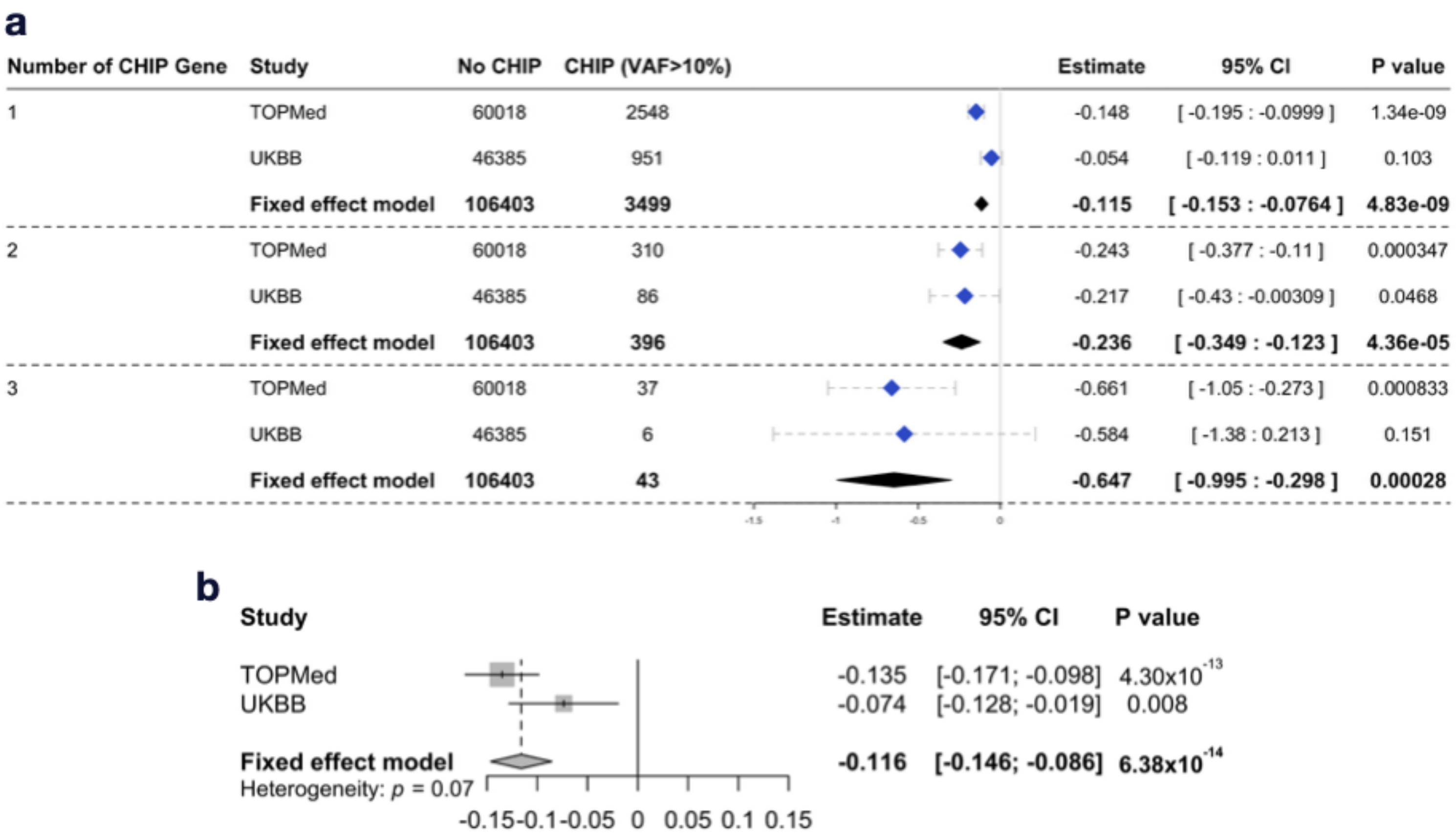


**a** The effect of CHIP on LTL was assessed by the number of CHIP related mutations. Linear regression model was adjusted with age, sex, smoking, BMI, first 11 genetic principal component, study, and sequencing center in both cohort and meta-analyzed using fixed effect model (study and sequencing center are only applicable to TOPMed). **b** The effect of the number of CHIP related mutations on LTL. CHIP: clonal hematopoiesis of indeterminate potential, CI: confidence interval, LTL: leukocyte telomere length, TOPMed: Trans-Omics for Precision Medicine, UKBB: UK Biobank, VAF: variant allele frequency.

**Supplementary Fig. 7: VAF is inversely correlated with LTL in both UK Biobank and TOPMed.**

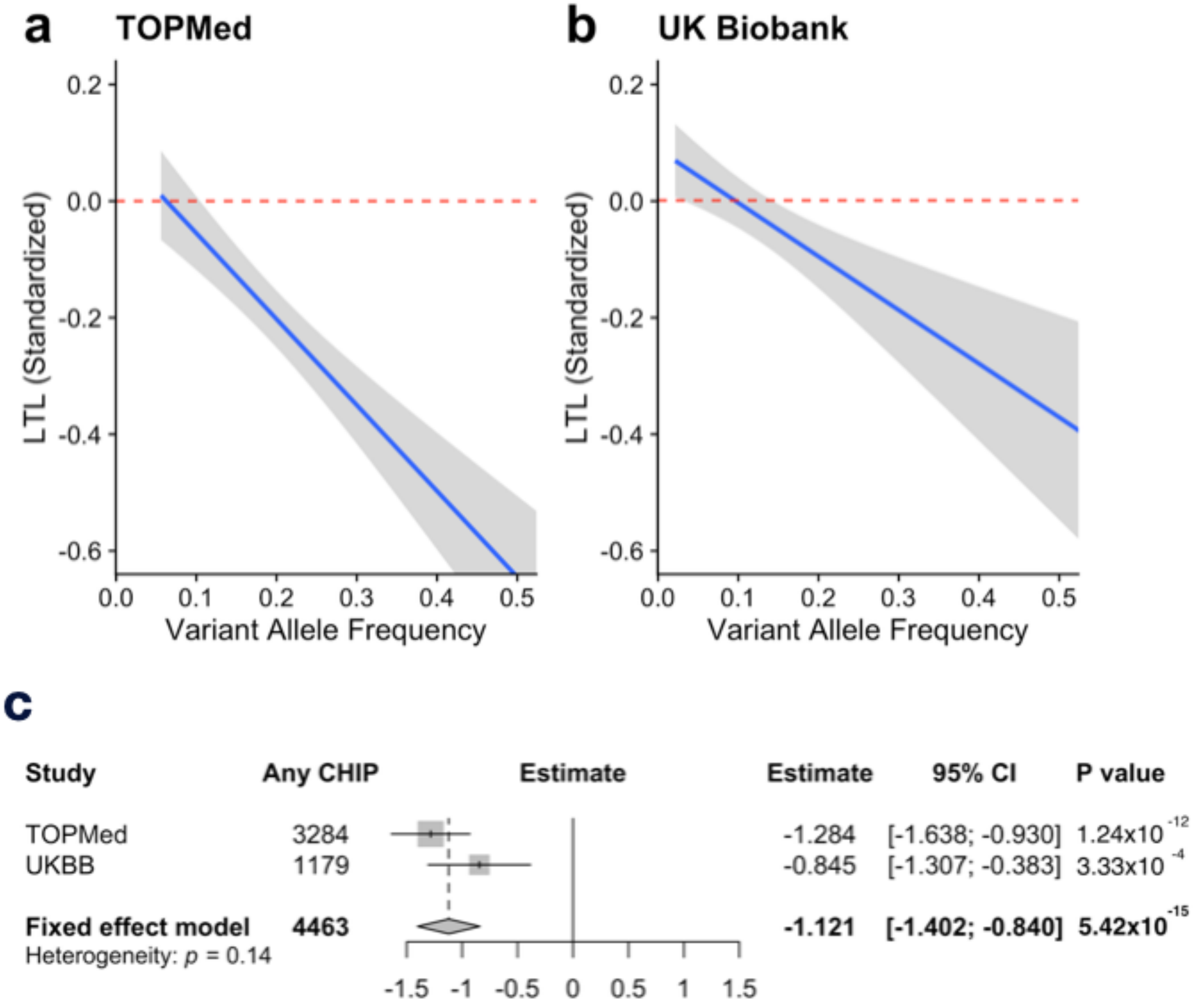


The association of VAF with LTL were assessed by linear regression model both in **a** TOPMed and **b** UK Biobank, then **c** meta-analyzed. Linear regression models were adjusted with age, sex, ever smoking, body mass index, first 11 principal components, study, and sequencing center (study and sequencing center are only applicable to TOPMed). CHIP: clonal hematopoiesis of indeterminate potential, CI: confidence interval, LTL: leukocyte telomere length, PC: principal component, TOPMed: Trans-Omics for Precision Medicine, UKBB: UK Biobank, VAF: variant allele frequency.

**Supplementary Fig. 8: Effect of shorter LTL on CAD.**

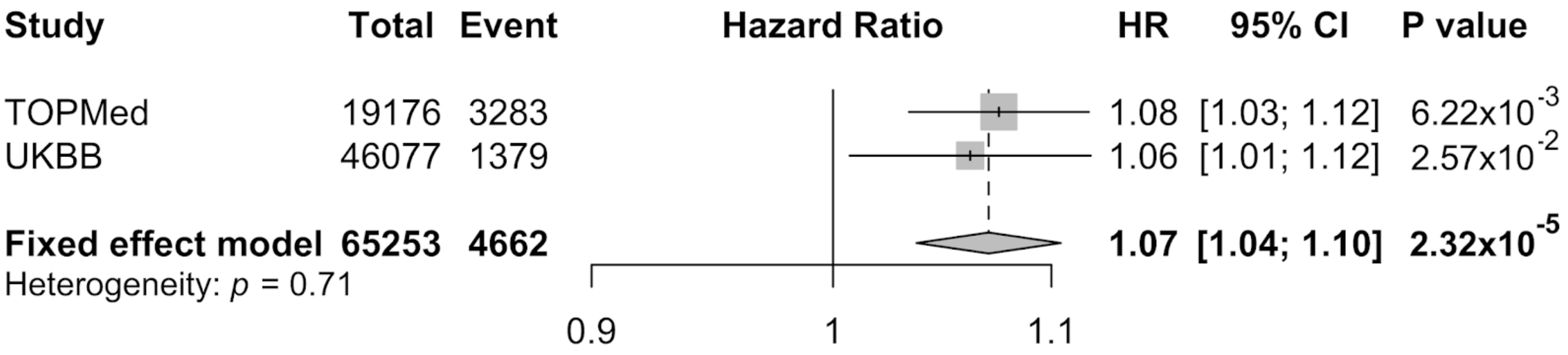


The effect of shorter LTL on CAD incidence was assessed in Cox Proportional Hazard model in both cohorts. The model was adjusted by age, sex, ever smoking, hypercholesterolemia, body mass index, first 11 genetic principal components. Effects were combined using fixed effects meta-analysis. CAD: coronary artery disease, HR: hazard ratio, TOPMed: Trans-Omics for Precision Medicine, UKBB: UK Biobank.

**Supplementary Fig. 9: Single instrumental variable tests of one-sample mendelian randomization for CHIP on LTL**

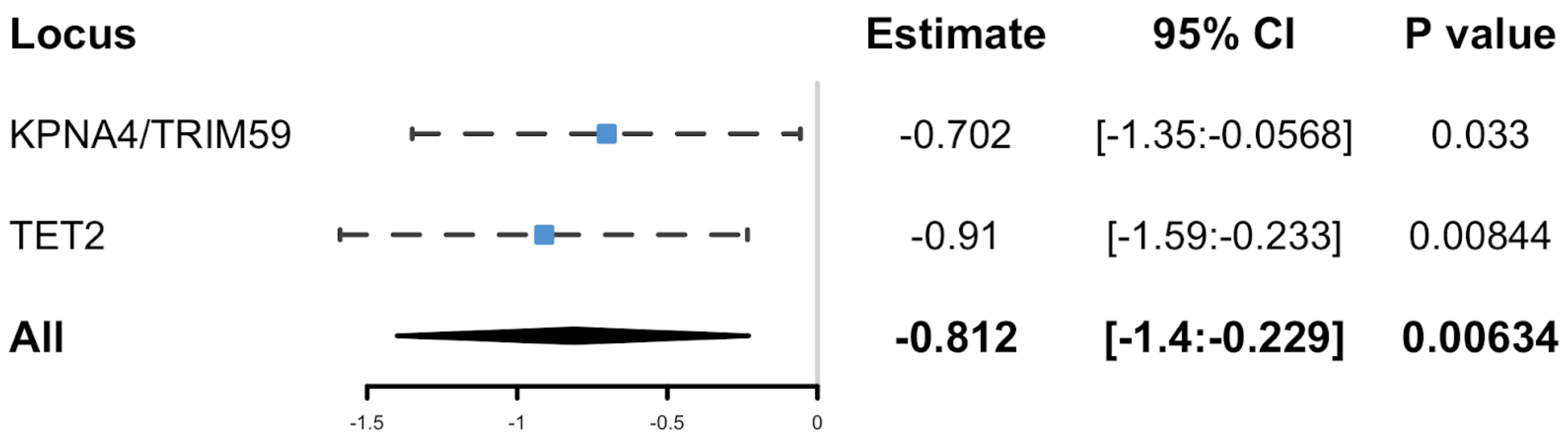


One-sample MR for CHIP on LTL using each variant separately. CHIP: clonal hematopoiesis of intermediate potential, CI: confidence interval, LTL: leukocyte telomere length.

**Supplementary Fig. 10: Leave-one-out analysis in MR-RAPS of CHIP on LTL.**

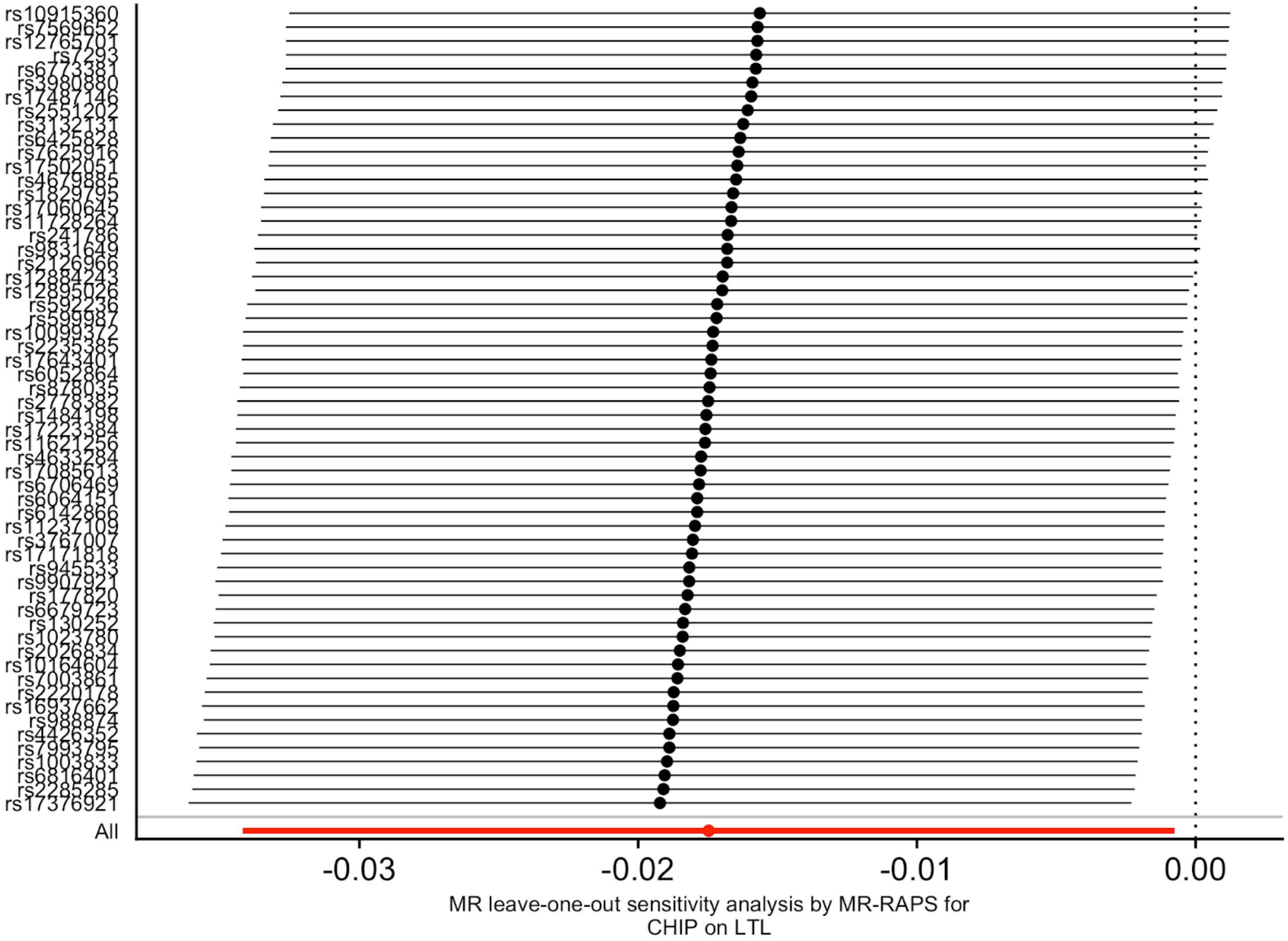


Leave-one-out analysis using MR-RAPS for CHIP on LTL. CHIP: clonal hematopoiesis of intermediate potential, LTL: leukocyte telomere length.

**Supplementary Fig. 11: Two-sample Mendelian randomization studies for LTL on CHIP.**

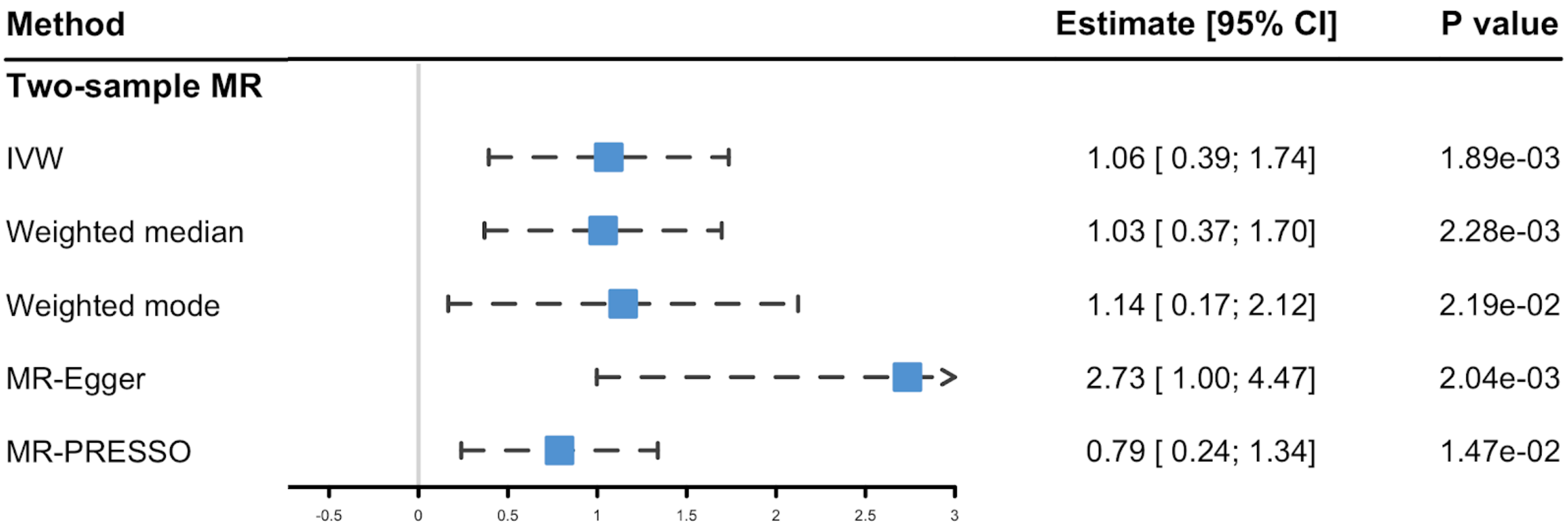


Two-sample MR studies for LTL on CHIP was performed to assess the causal effect of LTL on CHIP. IVs were derived Li et al *AJHG* 2020 in all analyses for LTL on CHIP and clumped as 10 Mb apart and in linkage disequilibrium ($R^2 > 0.001$ calculated in European ancestry from 1000 Genome project) resulting in 16 IVs. Li et al *AJHG* 2020 was used as the cohort of exposure and white British subset of UK Biobank was used for the outcome. In addition to the conventional inverse-variance weighted (IVW) method, weighted median, weighted mode, MR-Egger, and MR-PRESSO were performed as sensitivity analyses. MR-PRESSO excluded *TERT* and *ATM* loci variants as outliers. Used IVs and cohorts are reported in Supplementary Tables 6 and 7, respectively. CHIP: Clonal hematopoiesis of intermediate potential, CI: confidence interval, IV: Instrumental variable, LTL: Leukocyte telomere length, MR: Mendelian randomization, TOPMed: Trans-Omics for Precision Medicine, UKBB: UK Biobank.

**Supplementary Fig. 12: Leve-one-out analysis in two-sample MR of LTL on CHIP.**

| Excluded Variant | | β | 95% CI | P |
|---|---|---|---|---|
| **All Included** | | **1.06** | **[0.393:1.74]** | **0.00189** |
| rs4691895 | NAF1 | 1.19 | [0.545:1.83] | 0.000291 |
| rs228595 | ATM | 1.12 | [0.516:1.71] | 0.000265 |
| rs59294613 | POT1 | 1.09 | [0.41:1.76] | 0.00163 |
| rs3785074 | TERF2 | 1.06 | [0.384:1.74] | 0.00214 |
| rs2302588 | DCAF4 | 1.06 | [0.383:1.73] | 0.00214 |
| rs62053580 | RFWD3 | 1.04 | [0.356:1.73] | 0.00289 |
| rs10936600 | TERC | 1.03 | [0.27:1.79] | 0.00789 |
| rs9419958 | STN1(OBFC1) | 1.03 | [0.324:1.73] | 0.00423 |
| rs55749605 | SENP7 | 1.01 | [0.317:1.71] | 0.00433 |
| rs8105767 | ZNF208 | 0.99 | [0.296:1.68] | 0.00519 |
| rs2736176 | PRRC2A | 0.988 | [0.298:1.68] | 0.00501 |
| rs7194734 | MPHOSPH6 | 0.984 | [0.296:1.67] | 0.00507 |
| rs13137667 | MOB1B | 0.982 | [0.305:1.66] | 0.00449 |
| rs3219104 | PARP1 | 0.951 | [0.285:1.62] | 0.00509 |
| rs75691080 | RTEL1/STMN3 | 0.948 | [0.271:1.62] | 0.00607 |
| rs7705526 | TERT | 0.638 | [0.0224:1.25] | 0.0422 |

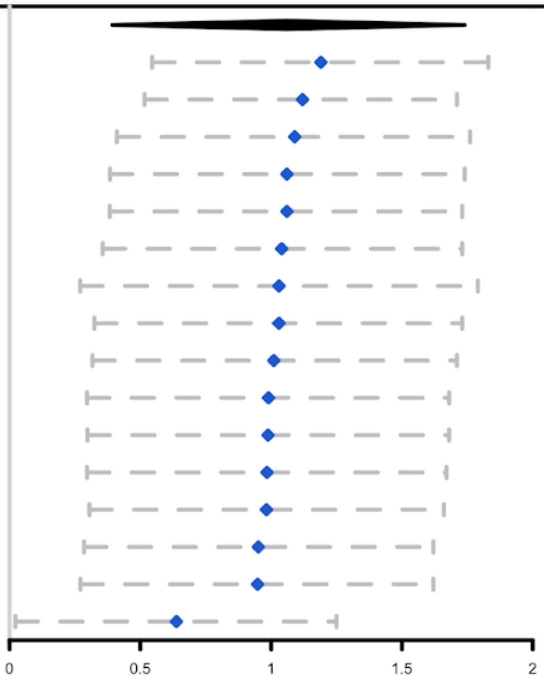


Leave-one-out analysis was performed by simple inverse variance weighted method. CHIP: clonal hematopoiesis of intermediate potential, LTL: leukocyte telomere length, MR: Mendelian randomization.

**Supplementary Fig. 13: Two-sample MR using 14 IVs excluding *TERT* and *ATM* loci**

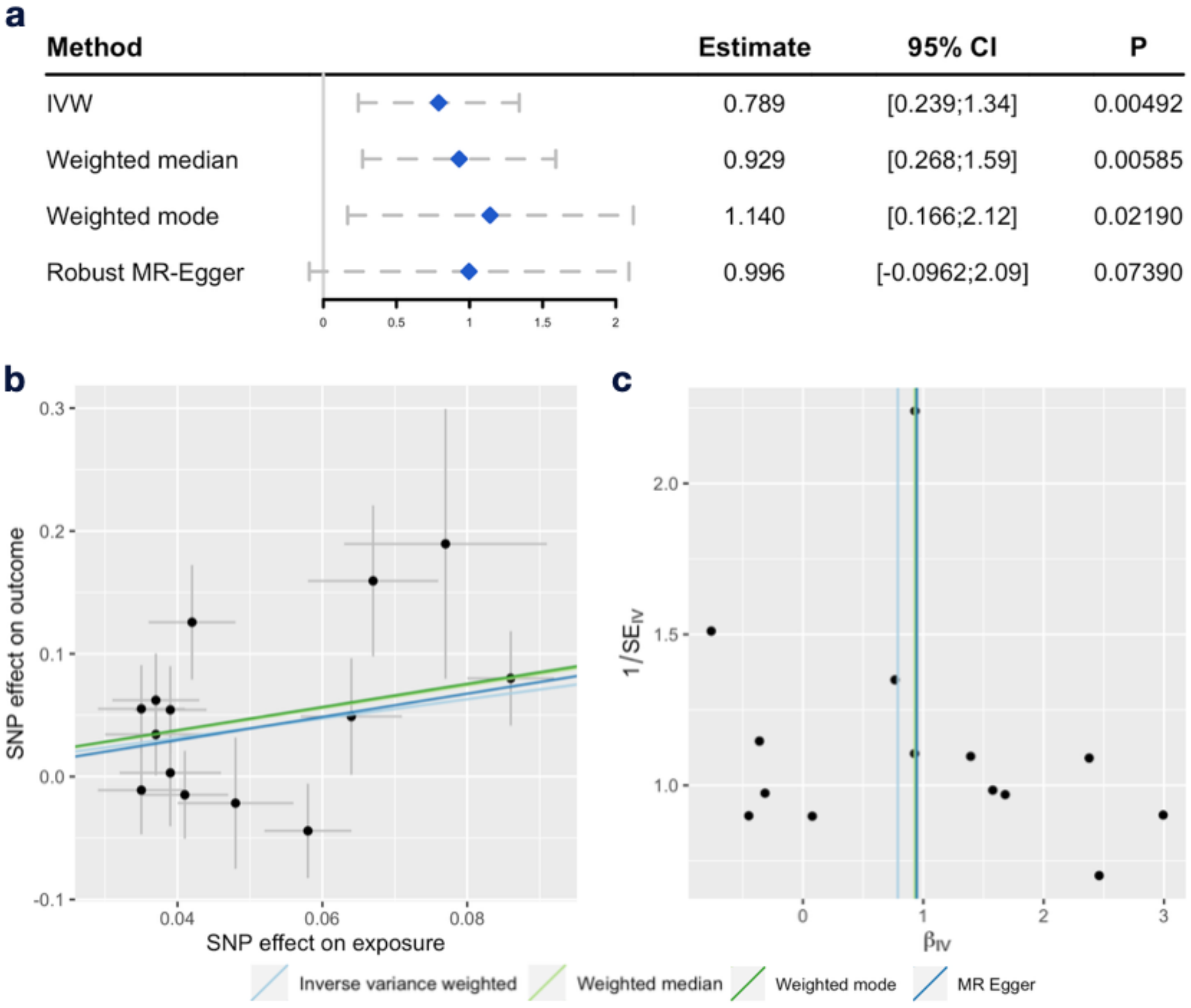


Two sample MR was performed using 14 IVs excluding outliers detected by MR-PRESSO. **a** Estimates from various methods using distinct assumptions. **b** Scatter plot and **c** Funnel plot for each IVs are displayed. IV: instrumental variables, IVW: inverse variance weighted, MR: Mendelian randomization

**Supplementary Fig. 14: Effect of LTL and genetic LTL score for mutation occurrence.**

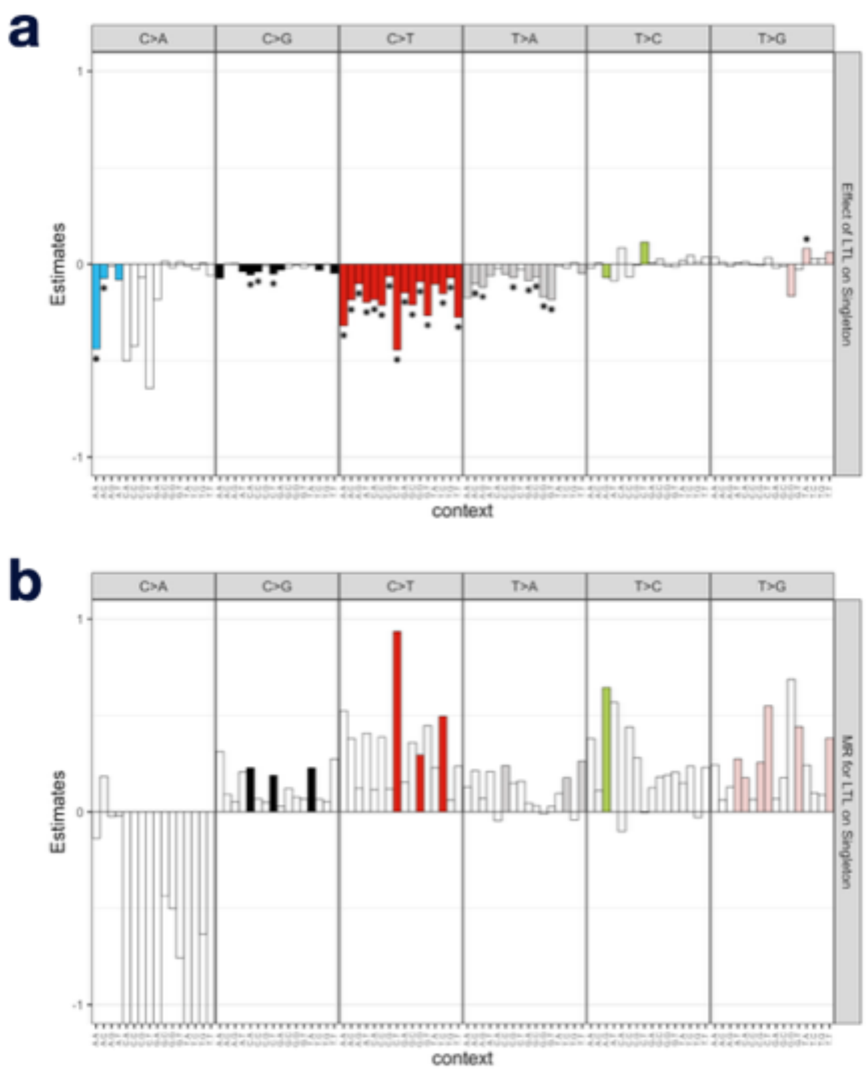


Effect estimates of **a** LTL and **b** one-sample MR using 14 IVs on singleton mutation occurrence. The vcf files were generated by Mutect2 from 49,953 CRAM files in TOPMed with appropriate filters and single base substitutions were extracted, stratified by trinucleotide context. IVs were selected as one-sample MR for LTL (Figure 3) with outlier exclusion. Effect estimates with $P < 0.05$ are colored. * denotes surviving from Bonferroni's correction. IV: instrumental variable, LTL: Leukocyte telomere length, MR: Mendelian randomization.

**Supplementary Fig. 15: Effect of LTL and genetic LTL score for COSMIC mutational signatures.**

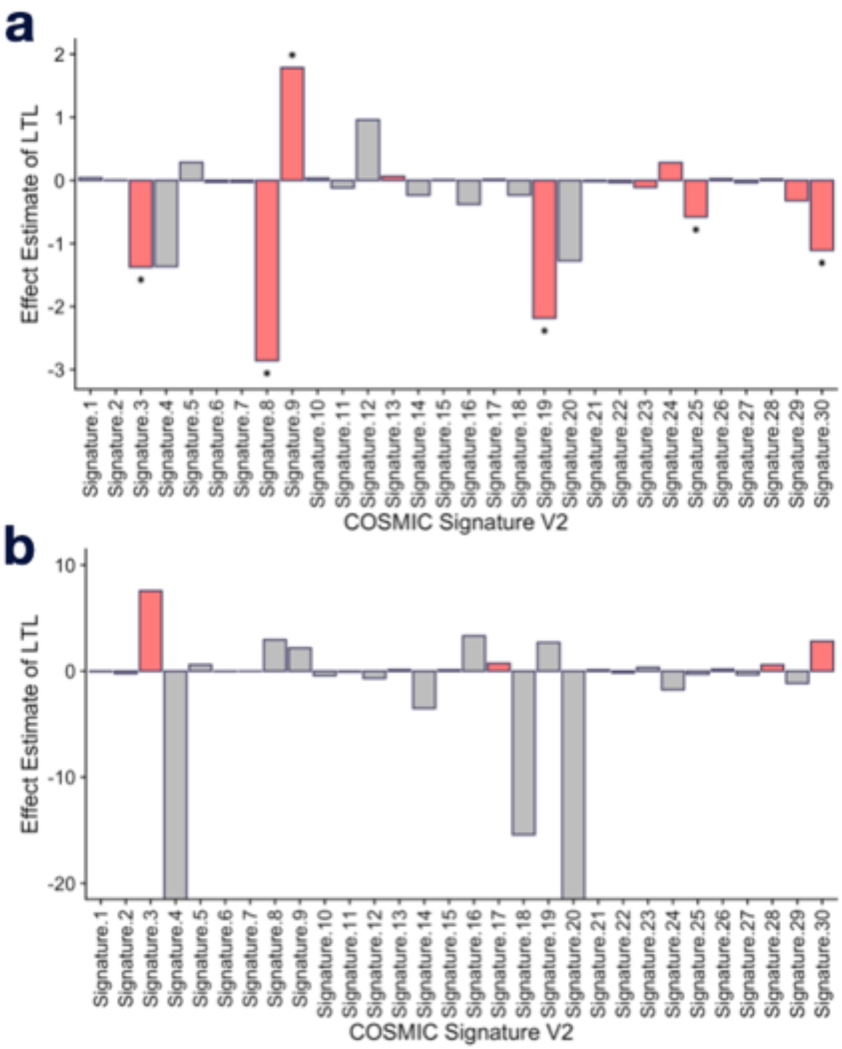


Effect estimates of **a** LTL and **b** one-sample MR for LTL on COSMIC mutational signature version 2 (https://cancer.sanger.ac.uk/cosmic/signatures_v2). *MutationalPattern* package in R was used to calculate the absolute contribution of each signature in each sample. The association with LTL was assessed by linear models. One-sample MR was performed by two-stage least-square method. Effect estimates with $P < 0.05$ are colored. * denotes surviving Bonferroni's correction. LTL: Leukocyte telomere length, MR: Mendelian randomization

**Supplementary Fig. 16: Effect of *TERT* locus for mutational occurrence and signature**

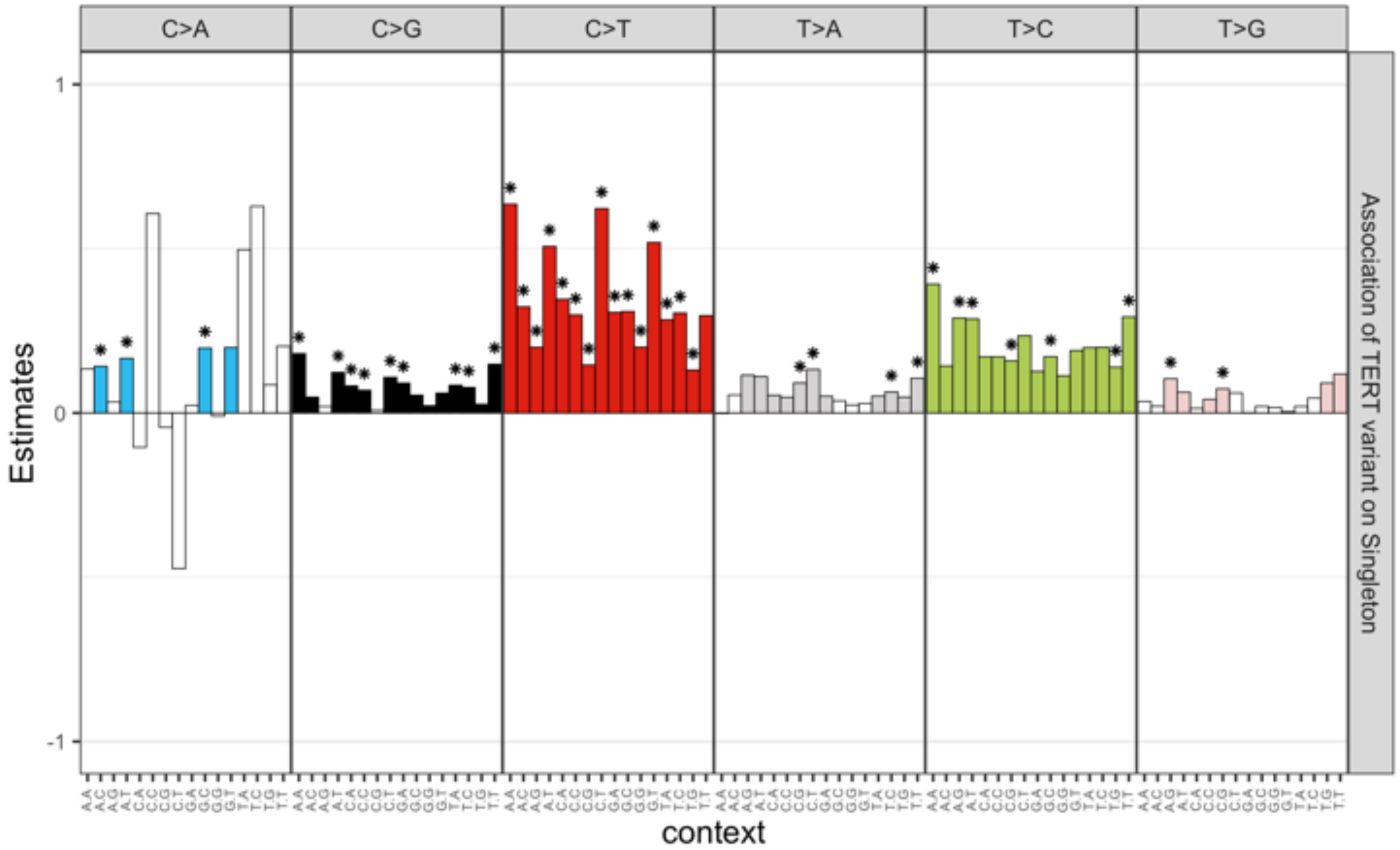


Effect estimates of *TERT* variant (rs7705526) on singleton mutation occurrence. The vcf files were generated by Mutect2 from 49,953 CRAM files in TOPMed with appropriate filters and single base substitutions were extracted, stratified by trinucleotide context. IVs were selected as one-sample MR for LTL (Figure 3) with outlier exclusion. Effect estimates with $P < 0.05$ are colored. * denotes surviving from Bonferroni's correction. LTL: Leukocyte telomere length, MR: Mendelian randomization

**Supplementary Fig. 17: Estimated change of mean LTL in each scenario.**

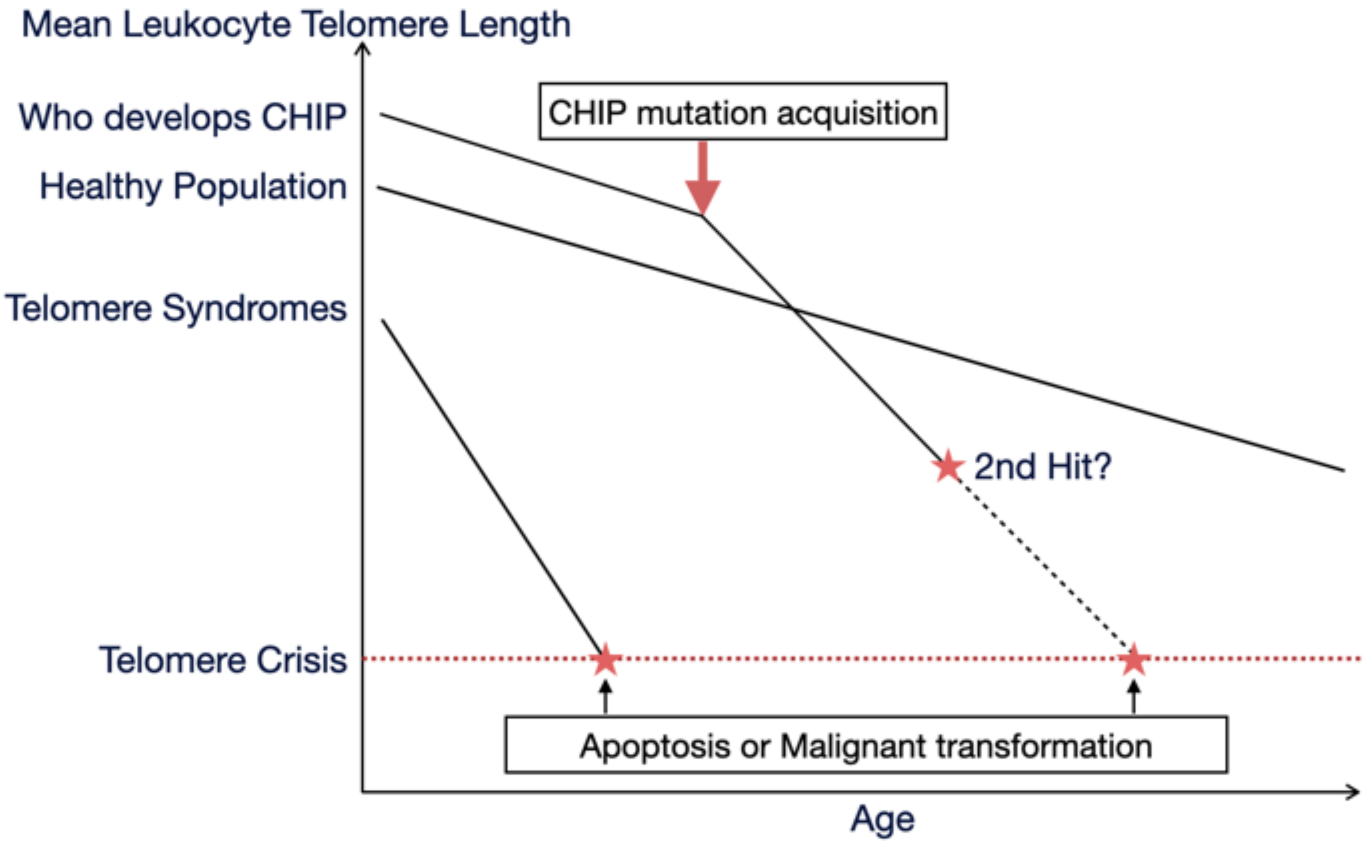


Schematic representation of estimated change of mean leukocyte telomere length in each scenario speculated from our study. CHIP: clonal hematopoiesis of intermediate potential, LTL: leukocyte telomere length.

**Supplementary Table 1: Baseline characteristics**

| | | TOPMed (N=63,302) | | | | UK Biobank (N=48,658) | | | |
|---|---|---|---|---|---|---|---|---|---|
| | | No CHIP (N=60,018) | CHIP VAF<0.1 (N=422) | CHIP VAF≧0.1 (N=2,862) | *P*-value | No CHIP (N=46,385) | CHIP VAF<0.1 (N=1,229) | CHIP VAF≧0.1 (N=1,044) | *P*-value |
| Age at blood draw (mean (SD)) | | 53.65 (18.12) | 66.50 (11.03) | 67.68 (10.64) | <0.001 | 56.36 (8.01) | 60.30 (6.65) | 60.80 (6.58) | <0.001 |
| Sex (Male, %) | | 25,581 (42.6) | 141 (33.4) | 1,073 (37.5) | <0.001 | 21,098 (45.5) | 542 (44.1) | 503 (48.2) | 0.136 |
| Race (%) | European | 29,171 (64.6) | 269 (73.7) | 1,854 (75.6) | <0.001 | 43,227 (93.9) | 1,160 (95.2) | 1,002 (96.3) | 0.013 |
| | African | 12,006 (26.6) | 76 (20.8) | 490 (20.0) | | 964 (2.1) | 20 (1.6) | 15 (1.4) | |
| | Asian | 2,665 (5.9) | 14 (3.8) | 69 (2.8) | | 1,019 (2.2) | 16 (1.3) | 11 (1.1) | |
| | Other | 1,347 (3.0) | 6 (1.6) | 40 (1.6) | | 829 (1.8) | 22 (1.8) | 13 (1.2) | |
| Smoking (%) | Never | 11,745 (19.6) | 113 (26.8) | 749 (26.2) | <0.001 | 25,857 (55.9) | 619 (50.7) | 494 (47.5) | <0.001 |
| | Previous | 40,089 (66.8) | 257 (60.9) | 1,738 (60.7) | | 16,241 (35.1) | 480 (39.3) | 435 (41.8) | |
| | Current | 8,184 (13.6) | 52 (12.3) | 375 (13.1) | | 4,136 (8.9) | 122 (10.0) | 112 (10.8) | |
| BMI, kg/m2 (mean (SD)) | | 28.55 (6.01) | 28.18 (5.74) | 28.24 (5.75) | 0.04 | 27.40 (4.79) | 27.52 (4.69) | 27.73 (4.77) | 0.075 |
| LTL, kb (mean (SD)) | Unadjusted | 3.28 (1.01) | 3.24 (0.98) | 3.14 (0.98) | <0.001 | 0.07 (0.08) | 0.07 (0.09) | 0.07 (0.08) | 0.658 |
| | Adjusted | 0.02 (1.00) | -0.18 (0.98) | -0.33 (0.97) | <0.001 | 0.00 (1.00) | 0.05 (1.02) | -0.12 (1.02) | <0.001 |
| Type 2 Diabetes (%) | | 1,946 (12.7) | 22 (18.2) | 111 (13.6) | 0.154 | 3,453 (7.4) | 114 (9.3) | 91 (8.7) | 0.018 |
| Hypercholesterolemia (%) | | 6,062 (10.1) | 74 (17.5) | 372 (13.0) | <0.001 | 10,884 (23.5) | 332 (27.0) | 315 (30.2) | <0.001 |

BMI: body mass index, CHIP: clonal hematopoiesis of intermediate potential, SD: standard deviation
TOPMed: Trans-Omics for Precision Medicine, VAF: variant allele frequecy.

**Supplementary Table 2: Effect estimates of each CHIP gene with VAF > 10 % on LTL**

| Gene | Study | n.control | n.case | Estimate | SE | *P*-value |
|---|---|---|---|---|---|---|
| *DNMT3A* | TOPMed | 60018 | 1496 | -0.019 | 0.031 | 0.544 |
| | UKBB | 46385 | 572 | -0.059 | 0.042 | 0.165 |
| | Fixed effect model | 106403 | 2068 | -0.033 | 0.025 | 0.190 |
| *TET2* | TOPMed | 60018 | 510 | -0.285 | 0.052 | 4.33E-08 |
| | UKBB | 46385 | 221 | -0.054 | 0.069 | 0.428 |
| | Fixed effect model | 106403 | 731 | -0.201 | 0.041 | 1.25E-06 |
| *ASXL1* | TOPMed | 60018 | 233 | -0.426 | 0.075 | 1.15E-08 |
| | UKBB | 46385 | 84 | -0.067 | 0.111 | 0.547 |
| | Fixed effect model | 106403 | 317 | -0.314 | 0.062 | 3.91E-07 |
| *PPM1D* | TOPMed | 60018 | 104 | -0.613 | 0.122 | 5.53E-07 |
| | UKBB | 46385 | 21 | -0.147 | 0.218 | 0.499 |
| | Fixed effect model | 106403 | 125 | -0.501 | 0.107 | 2.64E-06 |
| *JAK2* | TOPMed | 60018 | 92 | -0.798 | 0.130 | 8.93E-10 |
| | UKBB | 46385 | 9 | -0.197 | 0.332 | 0.554 |
| | Fixed effect model | 106403 | 101 | -0.718 | 0.121 | 3.17E-09 |
| *SF3B1* | TOPMed | 60018 | 72 | 0.004 | 0.135 | 0.978 |
| | UKBB | 46385 | 14 | -0.485 | 0.266 | 0.069 |
| | Fixed effect model | 106403 | 86 | -0.096 | 0.120 | 0.425 |
| *TP53* | TOPMed | 60018 | 54 | -0.665 | 0.173 | 1.21E-04 |
| | UKBB | 46385 | 21 | -0.395 | 0.223 | 0.076 |
| | Fixed effect model | 106403 | 75 | -0.563 | 0.137 | 3.72E-05 |
| *SRSF2* | TOPMed | 60018 | 50 | -0.035 | 0.166 | 0.833 |
| | UKBB | 46385 | 8 | -0.544 | 0.352 | 0.123 |
| | Fixed effect model | 106403 | 58 | -0.127 | 0.150 | 0.397 |
| *GNB1* | TOPMed | 60018 | 26 | -0.281 | 0.244 | 0.250 |
| | UKBB | 46385 | 11 | -0.302 | 0.301 | 0.314 |
| | Fixed effect model | 106403 | 37 | -0.290 | 0.190 | 0.127 |
| *GNAS* | TOPMed | 60018 | 22 | 0.076 | 0.279 | 0.787 |
| | UKBB | 46385 | 6 | 0.068 | 0.407 | 0.868 |
| | Fixed effect model | 106403 | 28 | 0.073 | 0.230 | 0.751 |
| *CBL* | TOPMed | 60018 | 17 | 0.248 | 0.279 | 0.375 |
| | UKBB | 46385 | 10 | -0.358 | 0.315 | 0.256 |
| | Fixed effect model | 106403 | 27 | -0.018 | 0.209 | 0.930 |
| *BRCC3* | TOPMed | 60018 | 16 | -0.128 | 0.260 | 0.621 |
| | UKBB | 46385 | 4 | 0.185 | 0.576 | 0.748 |
| | Fixed effect model | 106403 | 20 | -0.075 | 0.237 | 0.750 |
| *NF1* | TOPMed | 60018 | 13 | -0.807 | 0.411 | 0.050 |
| | UKBB | 46385 | 7 | 0.066 | 0.377 | 0.861 |
| | Fixed effect model | 106403 | 20 | -0.333 | 0.278 | 0.231 |
| *PRPF8* | TOPMed | 60018 | 15 | 0.046 | 0.381 | 0.905 |
| | UKBB | 46385 | 2 | 0.476 | 0.705 | 0.499 |
| | Fixed effect model | 106403 | 17 | 0.143 | 0.335 | 0.670 |
| *KRAS* | TOPMed | 60018 | 11 | 0.238 | 0.381 | 0.532 |
| | UKBB | 46385 | 4 | 0.721 | 0.498 | 0.148 |
| | Fixed effect model | 106403 | 15 | 0.416 | 0.302 | 0.169 |
| *ASXL2* | TOPMed | 60018 | 8 | -0.585 | 0.450 | 0.194 |
| | UKBB | 46385 | 4 | -0.264 | 0.499 | 0.597 |
| | Fixed effect model | 106403 | 12 | -0.441 | 0.334 | 0.187 |
| *CUX1* | TOPMed | 60018 | 6 | -0.281 | 0.712 | 0.693 |
| | UKBB | 46385 | 3 | 0.382 | 0.575 | 0.506 |
| | Fixed effect model | 106403 | 9 | 0.120 | 0.448 | 0.788 |

| | | | | | | |
|---|---|---|---|---|---|---|
| *IDH2* | TOPMed | 60018 | 6 | -0.159 | 0.450 | 0.724 |
| | UKBB | 46385 | 3 | 0.696 | 0.575 | 0.227 |
| | Fixed effect model | 106403 | 9 | 0.166 | 0.355 | 0.641 |
| *SETD2* | TOPMed | 60018 | 5 | -0.341 | 1.007 | 0.735 |
| | UKBB | 46385 | 4 | 0.091 | 0.498 | 0.855 |
| | Fixed effect model | 106403 | 9 | 0.006 | 0.447 | 0.990 |
| *BCORL1* | TOPMed | 60018 | 7 | -0.521 | 0.381 | 0.171 |
| | UKBB | 46385 | 1 | 1.859 | 0.997 | 0.062 |
| | Fixed effect model | 106403 | 8 | -0.218 | 0.356 | 0.540 |
| *BCOR* | TOPMed | 60018 | 6 | 0.095 | 0.581 | 0.870 |
| | UKBB | 46385 | 1 | 1.356 | 0.997 | 0.174 |
| | Fixed effect model | 106403 | 7 | 0.415 | 0.502 | 0.408 |
| *ETNK1* | TOPMed | 60018 | 5 | 0.136 | 0.503 | 0.786 |
| | UKBB | 46385 | 2 | -0.245 | 0.705 | 0.728 |
| | Fixed effect model | 106403 | 7 | 0.007 | 0.410 | 0.985 |
| *ETV6* | TOPMed | 60018 | 5 | 0.234 | 0.581 | 0.687 |
| | UKBB | 46385 | 2 | 0.310 | 0.705 | 0.660 |
| | Fixed effect model | 106403 | 7 | 0.265 | 0.448 | 0.555 |
| *RAD21* | TOPMed | 60018 | 5 | 0.100 | 0.712 | 0.889 |
| | UKBB | 46385 | 2 | -0.801 | 0.997 | 0.422 |
| | Fixed effect model | 106403 | 7 | -0.204 | 0.579 | 0.724 |
| *KDM6A* | TOPMed | 60018 | 4 | -1.204 | 0.581 | 0.038 |
| | UKBB | 46385 | 1 | -1.194 | 0.997 | 0.231 |
| | Fixed effect model | 106403 | 5 | -1.202 | 0.502 | 0.017 |
| *NRAS* | TOPMed | 60018 | 3 | 0.397 | 0.712 | 0.578 |
| | UKBB | 46385 | 2 | -0.765 | 0.705 | 0.278 |
| | Fixed effect model | 106403 | 5 | -0.190 | 0.501 | 0.704 |
| *SMC3* | TOPMed | 60018 | 2 | -0.946 | 0.712 | 0.184 |
| | UKBB | 46385 | 3 | 0.421 | 0.575 | 0.465 |
| | Fixed effect model | 106403 | 5 | -0.120 | 0.448 | 0.789 |
| *MPL* | TOPMed | 60018 | 2 | -2.680 | 1.007 | 0.008 |
| | UKBB | 46385 | 2 | -0.740 | 0.705 | 0.294 |
| | Fixed effect model | 106403 | 4 | -1.377 | 0.577 | 0.017 |
| *EP300* | TOPMed | 60018 | 2 | -2.572 | 1.007 | 0.011 |
| | UKBB | 46385 | 1 | 0.303 | 0.997 | 0.761 |
| | Fixed effect model | 106403 | 3 | -1.120 | 0.708 | 0.114 |
| *SUZ12* | TOPMed | 60018 | 2 | -0.058 | 0.712 | 0.935 |
| | UKBB | 46385 | 1 | 0.640 | 0.997 | 0.521 |
| | Fixed effect model | 106403 | 3 | 0.178 | 0.579 | 0.759 |
| *BRAF* | TOPMed | 60018 | 1 | 1.660 | 1.007 | 0.099 |
| | UKBB | 46385 | 1 | 0.005 | 0.997 | 0.996 |
| | Fixed effect model | 106403 | 2 | 0.824 | 0.708 | 0.245 |
| *SMC3* | TOPMed | 60018 | 2 | -0.943 | 0.712 | 0.185 |
| | UKBB | 46117 | 4 | -0.712 | 0.498 | 0.153 |
| | Fixed effect model | 106135 | 6 | -0.788 | 0.408 | 0.054 |
| *NRAS* | TOPMed | 60018 | 3 | 0.387 | 0.712 | 0.587 |
| | UKBB | 46117 | 2 | -0.773 | 0.705 | 0.273 |
| | Fixed effect model | 106135 | 5 | -0.199 | 0.501 | 0.691 |

CHIP: clonal hematopoiesis of intermediate potential, SE: standard error
TOPMed: Trans-Omics for Precision Medicine, UKBB: UK Biobank, VAF: variant allele frequecy.

**Supplementary Table 3: Coronary artery disease definition in UK Biobank.**

| Dianosis | Disease | Code |
| --- | --- | --- |
| Self reported | Heart attack/Myocardial infarction | 1075 |
| | Coronary angioplasty/bypass grafting | 1070, 1095, 1523 |
| Diagnosed by doctor | Heart attack | 1 |
| ICD9 | Myocardial infarction | 410, 4109, 412, 4129 |
| | Other ischemic heart disease | 411, 4119 |
| ICD10 | Myocardial infarction | I21, I21.0, I21.1, I21.2, I21.3, I21.4, I21.9, I22, I22.0, I22.1, I22.8, I22.9, I23, I23.0, I23.1, I23.2, I23.3, I23.4, I23.5, I23.6, I23.8, I24.0, I24.1, I25.2 |
| | Other ischemic heart disease | I24, I24.8, I24.9 |
| OPCS | Coronary angioplasty/bypass grafting | K40, K40.1, K40.2, K40.3, K40.4, K40.8, K40.9, K41, K41.1, K41.2, K41.3, K41.4, K41.8, K41.9, K42, K42.1, K42.2, K42.3, K42.4, K42.8, K42.9, K43, K43.1, K43.2, K43.3, K43.4, K43.8, K43.9, K44, K44.1, K44.2, K44.8, K44.9, K45.1, K45.2, K45.3, K45.4, K45.5, K45.6, K45.8, K45.9, K46, K46.1, K46.2, K46.3, K46.4, K46.5, K46.8, K46.9, K49.1, K49.2, K49.3, K49.4, K49.8, K49.9, K50.1, K50.2, K50.4, K75.1, K75.2, K75.3, K75.4, K75.8, K75.9 |

**Supplementary Table 5: Instrumental variables used in one-sample Mendelian randomization of CHIP on LTL**

| SNP ID | Chromosome | Position | Gene | F value for CHIP | R square for CHIP | R square for LTL | Steiger test | | |
|---|---|---|---|---|---|---|---|---|---|
| | | | | | | | Steiger test Direction | t value | *P*-value |
| rs58322641 | 3 | 160497760 | *KPNA4/TRIM59* | 38.681 | 9.50E-04 | 7.06E-05 | TRUE | 4.68 | 2.90E-06 |
| rs114420266 | 4 | 104838707 | *TET2* | 36.303 | 1.62E-04 | 8.23E-06 | TRUE | 2.06 | 0.0399 |
| rs7705526 | 5 | 1285859 | *TERT* | | 0.00124 | 0.00225 | FALSE | -2.56 | 0.0105 |

CHIP: clonal hematopoiesis of intermediate potential
LTL: leukocyte telomere length.

**Supplementary Table 6: Instrumental variables used for two-sample MR study for CHIP on LTL.**

| rsid | Effect Allele | Chromosome | Position (GRCh38) |
|---|---|---|---|
| rs10915360 | C | 1 | 4057494 |
| rs4633284 | G | 1 | 15006557 |
| rs6425828 | G | 1 | 33576217 |
| rs599987 | A | 1 | 78130644 |
| rs2026834 | A | 1 | 114188884 |
| rs3767007 | T | 1 | 181752820 |
| rs6679723 | C | 1 | 240046393 |
| rs7569652 | C | 2 | 52616205 |
| rs10164604 | A | 2 | 134296795 |
| rs2551202 | C | 2 | 221826117 |
| rs6706469 | T | 2 | 239472917 |
| rs6773381 | G | 3 | 29211616 |
| rs7625916 | G | 3 | 113601212 |
| rs4679885 | G | 3 | 160436959 |
| rs9831649 | G | 3 | 166899039 |
| rs6816401 | T | 4 | 14360947 |
| rs11728264 | T | 4 | 29775960 |
| rs17085613 | T | 4 | 55313200 |
| rs2220178 | C | 4 | 93485971 |
| rs1829795 | C | 4 | 180960087 |
| rs2126966 | C | 5 | 25470572 |
| rs17171818 | T | 5 | 138389314 |
| rs2235385 | T | 6 | 11775488 |
| rs3132131 | G | 6 | 32931708 |
| rs592236 | C | 6 | 140855084 |
| rs17502051 | G | 7 | 27265763 |
| rs3980880 | A | 7 | 67286012 |
| rs17487146 | G | 7 | 147193170 |
| rs2285285 | A | 8 | 17341221 |
| rs10099372 | G | 8 | 30567398 |
| rs16937662 | A | 8 | 71568209 |
| rs7003861 | C | 8 | 124020492 |
| rs2778382 | A | 9 | 85845149 |
| rs17376921 | C | 10 | 11135204 |
| rs988874 | A | 10 | 27395731 |
| rs2675592 | T | 10 | 61851775 |
| rs12765701 | T | 10 | 114500429 |
| rs11237109 | G | 11 | 77191550 |
| rs945533 | A | 13 | 91553728 |
| rs878035 | A | 13 | 106426937 |
| rs7993795 | T | 13 | 107681001 |
| rs12895026 | A | 14 | 58533544 |
| rs12884243 | G | 14 | 76825609 |
| rs11621256 | T | 14 | 98955293 |
| rs1484198 | T | 15 | 46385075 |
| rs1023780 | T | 15 | 98289869 |
| rs4426352 | A | 16 | 26592687 |
| rs17643401 | A | 17 | 7601540 |
| rs1003833 | G | 17 | 13336508 |
| rs241786 | C | 17 | 28239010 |
| rs9907921 | T | 17 | 76260832 |
| rs17060645 | A | 18 | 77199324 |
| rs177820 | G | 18 | 79516964 |
| rs6052864 | G | 20 | 4771533 |
| rs6064151 | A | 20 | 55018501 |
| rs6142866 | C | 20 | 61793530 |
| rs130252 | C | 22 | 33343549 |
| rs7293 | T | 22 | 35611028 |
| rs17223384 | G | 22 | 48647820 |

CHIP: clonal hematopoiesis of intermediate potential
LTL: leukocyte telomere length
MR: Mendelian randomization

**Supplementary Table 7: Instrumental variable used in Mendelian randomization studies for LTL on CHIP.**

| rsid | Gene | Chromosome | Position (GRCh38) | RSSobs | *P*-value |
|---|---|---|---|---|---|
| rs3219104 | PARP1 | 1 | 226374920 | 1.40E-05 | 0.6432 |
| rs55749605 | SENP7 | 3 | 101513249 | 2.47E-07 | 1 |
| rs10936600 | TERC | 3 | 169796797 | 8.29E-07 | 1 |
| rs13137667 | MOB1B | 4 | 70908630 | 1.75E-05 | 1 |
| rs4691895 | NAF1 | 4 | 163127047 | 2.22E-05 | 0.0816 |
| rs7705526 | TERT | 5 | 1285859 | 4.90E-05 | <0.0016 |
| rs2736176 | PRRC2A | 6 | 31619784 | 2.22E-06 | 1 |
| rs59294613 | POT1 | 7 | 124914213 | 1.12E-05 | 0.4656 |
| rs9419958 | STN1(OBFC1) | 10 | 103916188 | 8.97E-07 | 1 |
| rs228595 | ATM | 11 | 108234866 | 2.45E-05 | 0.0048 |
| rs2302588 | DCAF4 | 14 | 72938044 | 5.82E-06 | 1 |
| rs3785074 | TERF2 | 16 | 69373083 | 4.78E-06 | 1 |
| rs62053580 | RFWD3 | 16 | 74646176 | 8.79E-06 | 1 |
| rs7194734 | MPHOSPH6 | 16 | 82166375 | 6.10E-07 | 1 |
| rs8105767 | ZNF208 | 19 | 22032639 | 2.43E-07 | 1 |
| rs75691080 | RTEL1/STMN3 | 20 | 63638397 | 7.38E-06 | 1 |

RSSobs: Observed residdual sum of squares calculated by MR-PRESSO.

*P*-value: Heterogeneity test assessed by MR-PRESSO

**Supplementary Table 8: Cohorts used for Mendelian randomization studies**

| | | IV discovery | Exposure | Outcome |
|---|---|---|---|---|
| CHIP on LTL | One-sample MR | TOPMed | | |
| | Two-sample MR | TOPMed (European ancestry) | | ENGAGE |
| LTL on CHIP | One-sample MR | Li et al, *AJHG* 2020 | TOPMed | |
| | Two-sample MR | Li et al, *AJHG* 2020 | | UK biobank (White British) |

*AJHG*: American Journal of Human Genetics
CHIP: clonal hematopoiesis of intermediate potential
ENGAGE: European Network for Genetic and Genomic Epidemiology
LTL: leukocyte telomere length
MR: Mendelian randomization
TOPMed: Trans-Omics for Precision Medicine
